# Health Care Workload Impacts and Cost-Effectiveness of a Metabolomic Risk Score-based Health Check for Cardiometabolic Disease Prevention in Finland

**DOI:** 10.1101/2025.09.11.25335561

**Authors:** Piia Lavikainen, Leena Haikonen-Salo, Aku-Ville Lehtimäki, Kari Jalkanen, Jari Heiskanen, Tiina Laatikainen, Janne Martikainen

## Abstract

**Background:** Predictive metabolomics enables efficient early cardiometabolic risk detection, supporting proactive prevention over traditional reactive healthcare approaches. We evaluated the impact of a novel metabolomic risk score (MRS)-based health check on healthcare workload and long-term cost-effectiveness compared with current practices in Finland’s working-age population.

**Methods:** A de novo individual-level microsimulation model was developed to estimate labour time and cost-effectiveness when identifying individuals at risk of cardiometabolic diseases (cardiovascular diseases; CVD or type 2 diabetes; T2D). The model used synthetic data on 256,372 Finnish individuals aged 50–54 years without prior CVD or T2D. From a societal perspective, three scenarios were assessed: 1) replacing the standard health checks with MRS-based health checks, 2) replacing standard health checks with MRS-based checks plus enhanced prevention, and 3) comparing enhanced standard checks with MRS-based checks plus enhanced prevention. Outcomes included time required to identify at-risk individuals, incremental cost-effectiveness ratio per QALY gained, and cost-effectiveness acceptability curves.

**Results:** MRS-based health checks substantially reduce workload, saving 193,140 hours for nurses and 2,843 hours for physicians over five years. The approach is likely cost-saving across all scenarios, with discounted long-term savings ranging from €27 million to €258 million. In scenarios 1–2, QALYs improve, with gains from 1,316 to 10,813. In scenario 3, no QALY gains are observed, and minor losses occur due to differences in baseline risk stratification.

**Conclusions:** MRS-based health checks in primary and occupational care reduce workload and are likely a cost-saving strategy with potential health outcome benefits in identifying individuals at risk of cardiometabolic diseases.

**Plain language summary:** Heart disease, stroke, and type 2 diabetes are common and costly health problems. Finding people at risk early can help prevent these diseases. This study looked at a new blood-test-based health check and compared it with current health checks in Finland. Researchers used a computer model based on over 250,000 people aged 50–54. They tested different ways of using the new test and compared time, costs, and health outcomes. The results suggested that the new test can save time for nurses and physicians and reduce healthcare costs. In most cases, it also improved health outcomes. This means that using this type of test could help healthcare services work more efficiently and support earlier prevention, which may benefit both individuals and society in the future.

**Funding:** This work was supported by Nightingale Health Plc.

## Introduction

Cardiometabolic diseases (CMD), such as coronary heart disease (CHD), stroke, and type 2 diabetes (T2D), represent one of the major health and economic challenges in the world^1,2^. In Europe alone, cardiovascular diseases (CVDs) cause around 32% of deaths annually^3^. These deaths and CVD-related morbidity together impose an annual economic burden estimated at 282 billion euros within the European Union^4^. In addition, due to increase in the prevalence of obesity caused by unfavourable trends in excess energy intake, unhealthy dietary habits, and low physical activity, there are about 60 million people with T2D in Europe, a condition known to significantly increase the risk of macrovascular (i.e. 1.5- to 2-fold higher risk for CVDs)^5,6^ and microvascular complications such as retinopathy, neuropathy, and nephropathy^7^. CVDs are the main cause of death in patients with T2D^8,9^.

Traditionally, healthcare systems have primarily focused on the reactive treatment of CMDs (i.e., addressing them only after they arise). However, accumulating evidence underscores the necessity of shifting towards proactive prevention of CMDs, emphasizing early risk identification, lifestyle interventions, and optimized management of modifiable risk factors such as dyslipidaemia, hypertension, and obesity^10,11^. This paradigm shift is essential not only for reducing the incidence and severity of CMDs but also for alleviating healthcare costs and improving long-term health outcomes. Unfortunately, one of the primary obstacles in proactive prevention is the lack of resources necessary for early detection and preventive interventions for these conditions through comprehensive population-wide health checks. Although comprehensive population-wide health checks have been widely implemented in some countries, evidence on their cost effectiveness remains mixed. Large systematic reviews^12^ have shown little or no impact on mortality outcomes, which partly explains why such programmes are not universally prioritised under constrained health care resources. More recent evidence suggests that targeted health checks may be more cost effective than universal approaches^13^. This highlights the need for more scalable, stratified, engaging, and cost-effective strategies to improve early detection and prevention of CMD in a precise manner^14^.

Currently, predictive metabolomics is increasingly used in identifying individuals at high risk for many chronic diseases^15^. Recent scientific developments have enabled the analysis of blood samples and other risk factors, providing detailed insights into an individual’s metabolic profile and its potential associations with the future disease risks^15,16^. Predictive metabolomics using high-throughput nuclear magnetic resonance (NMR) spectroscopy, such as the Nightingale Health platform, enables the quantification of a standardized panel of circulating metabolic biomarkers, including detailed lipoprotein subclasses. These measures have been combined with large longitudinal biobank datasets to develop population-level risk prediction models, typically based on regression approaches, for a wide range of diseases^15,16^. These models consider the individual’s metabolic profile and other relevant factors to estimate their likelihood of developing different diseases in the future. Individuals at high risk can be prioritized for more rigorous preventive measures in line with clinical guidelines, thereby reducing their disease risk.

Thus, by accurately pinpointing individuals at elevated risk for diseases healthcare systems can allocate their limited resources more efficiently. Furthermore, the developed approach offers significant potential to streamline workflows and reduce workload in healthcare by simplifying risk assessments. Large population-based cohort studies have shown that metabolomic risk scores (MRSs) improve risk discrimination and reclassification beyond established clinical prediction models such as SCORE2, with modest but population relevant gains in C statistics and net reclassification indices^17^. However, to our knowledge, there is no prior evidence on the economic implications of predictive metabolomics.

Since MRS can be measured from a standard blood sample, it enables feasible risk assessments in today’s cost-constrained health systems by reducing the time healthcare professionals spend on current risk assessment procedures. Moreover, predictive metabolomics can pave the way for targeted preventive therapies, ultimately leading to improved population health outcomes and reduced healthcare costs in the long run. However, due to above mentioned limited health care resources, careful considerations of these potential workforce impacts, and wider health economic consequences of novel risk assessment tools and interventions are needed to support the informed decision-making on their large-scale implementation and utilization to rationalize resource allocation in health care. In the present study, we focused on to assess the health care professionals’ workload impacts and long-term cost-effectiveness of a novel MRS-based health checks in a general working-age Finnish population.

This study demonstrates that MRS-based health checks reduce healthcare workload and are cost-saving across all modelled scenarios, with discounted long-term savings ranging from €27 million to €258 million and potential quality-adjusted life year gains in scenarios where at-risk individuals are identified and treated.

## Methods

A *de novo* individual-level microsimulation model was developed to assess the expected changes in health care professionals’ labour time when conducting health checks and the long-term cost-effectiveness of the MRS-based health check program among the general working-age adult population, compared with current health check practices in Finland. The general workflow of the model development is illustrated in Fig. 1.

**Fig. 1.**
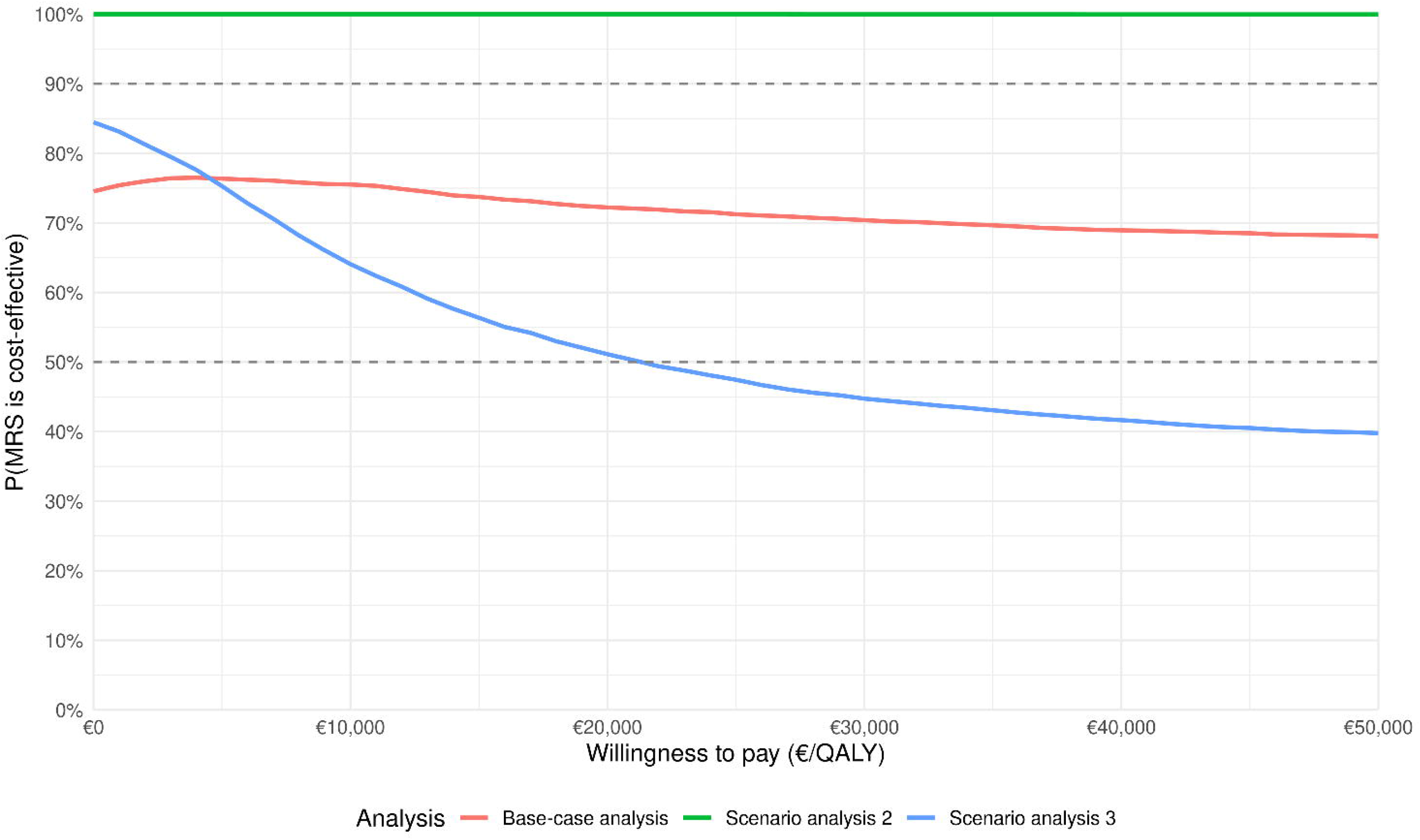
General workflow of the health economic model development. The THL Biobank dataset was used to develop a synthetic target population and parametric disease-free survival models, which informed the development and validation of a microsimulation health economic model. Three health check scenarios comparing MRS-based and standard health checks, with and without enhanced prevention, were evaluated for their impact on healthcare professionals’ workload, long-term costs, and health outcomes including QALYs. *Finnish individuals aged 50–54 years with no prior history of T2D or cardiovascular disease, representing a working-age subpopulation approaching the age at which cardiometabolic disease incidence typically begins to rise. Abbreviations: CHD, coronary heart disease; HC, health check; HDL, high-density lipoprotein cholesterol; MRS, metabolic risk score; QALY, quality-adjusted life year; SBP, systolic blood pressure; T2D, type 2 diabetes; TC, total cholesterol.

The simulation model was populated with synthetic (hypothetical) Finnish individuals aged 50–54 years who had no history of T2D or CVD; representing a working-age subpopulation approaching the age when the sharpest increase of CMD is usually observed^18^. In 2023, the total population within this age group was 315,799 individuals in Finland^19^. Of this population, 89% were part of the labour force^20^, and 9.3% had prevalent CVD or T2D^21^. Assuming that 4.7% of individuals with CHD and 24.2% of those with a history of stroke were not in the labour force^22,23^, the proportion of the labour force with CVD or T2D was estimated at 8.8%. Therefore, the final size of the target population amounted to 256,372 individuals.

To compare the MRS-based health check against the standard health check, two alternative study arms were modelled. In the standard health check arm, all individuals in the target population were assumed to be invited to one health check during the five-year implementation period. Individuals’ attendance was based on current Finnish practice. Annually, 15.6% of employed individuals attended occupational health nurse checks^24,25^, while 8.6% of unemployed individuals attended public health checks^26^. Given an unemployment rate of 5.3% for the target age group in 2023^27^, the overall annual attendance rate was calculated as a weighted average: 15.6% × (100% − 5.3%) + 8.6% × 5.3% = 15.2% (see Supplementary File section 3.1 for details). In the MRS-based health check arm, all individuals were invited for MRS risk testing (Nightingale Health Plc risk scores are currently utilized in Finnish health care settings^28,29^; methodology described in Nightingale Health Biobank Collaborative Group^16^) in the first year, with an assumed attendance rate equivalent to that of standard health checks (i.e., 76.0% over five years), but concentrated within the first year of the model because saved resources enables more efficient risk identification.

In the standard health check arm, individuals attended conventional health checks, including a laboratory visit and a nurse visit. During the nurse visit, moderate- and high-risk individuals were identified according to the Finnish Current Care Guidelines using FINDRISC and FINRISK risk scores^18,30^ as part of the standard health check process in Finland^31^. In addition to laboratory measurements, the determination of both the scores requires collection of basic clinical and anthropometric data during the nurse’s appointment, including measurements of height, weight, and waist circumference, as well as assessment of blood pressure and structured information on lifestyle and medical history obtained through a standardized questionnaire. In the MRS-based health check arm, individuals were assumed to undergo MRS testing instead of conventional health checks (see Supplementary File section 3.2 for details), requiring only a laboratory visit. Nightingale MRS was used (see Supplementary File section 2.1.2). MRS was derived from metabolomic biomarkers measured using a nuclear magnetic resonance spectroscopy based platform that quantifies lipoprotein particles, lipid fractions, and low molecular weight metabolites relevant to CVD and T2D^15,16^. Participants were classified into three risk categories (high, moderate, and low) based on their estimated CVD and T2D risk, with classification determined by the higher risk score of the two conditions. Both in the standard health check arm and in the MRS-based health check arm, individuals at high risk were referred to a physician visit. Thus, there was a difference in workload between the study arms: the nurse visit required to identify risk in the standard health check arm was not needed in the MRS-based health check.

In both arms, the underlying “true” risks of CMD were estimated using parametric survival regression models incorporating both MRS-based and conventional risk factors. The use of both risk factor types was expected to maximize the predictive accuracy of disease risk across different risk categories. Preventive interventions for individuals at moderate and high risk were assumed to be identical in both study arms. To assess the added value of conventional and MRS-based health checks combined with enhanced preventive measures, scenario analyses were conducted also for optimized interventions. These interventions incorporated stricter thresholds for high blood pressure and hyperlipidemia, along with additional measures targeting overweight, obesity, and unhealthy diet.

A recently introduced Time Needed to Treat (TNT) approach^32^ was applied to evaluate potential shifts in healthcare professionals’ labour time requirements between the study arms. The TNT approach was used to estimate the time required to identify a single individual at elevated risk for CMD under the conditions of each arm. It was assumed that, per health check, a nurse would spend approximately one hour, a physician 30 minutes, and a laboratory professional 10 minutes (see Supplementary File section 4.1 for details). For both nurse and physician visits, the estimated time included the full scope of the appointment: the reception, patient consultation and counselling, as well as documentation and other related administrative tasks^33^. MRS testing was estimated to require only 10 minutes of laboratory professional time per individual and administrative tasks were assumed to be integrated into existing digital systems and therefore required minimal additional healthcare personnel time compared with the standard health check.

To assess the long-term cost-effectiveness of MRS-based health check, health state-specific costs and quality-of-life weights were sourced from the literature to estimate cumulative long-term costs and quality-adjusted life years (QALYs). These estimates were used to calculate the incremental cost-effectiveness ratio (ICER) per QALY gained between the study arms. The analysis was conducted from a societal perspective, incorporating both direct costs and productivity loss costs. In the base-case analysis, a lifetime horizon and a 3% annual discount rate for costs and QALYs were applied in accordance with national guidelines for health economic evaluations^34,35^. All costs were adjusted to 2022 levels using the official price index of public expenditure and the index of wage and salary earnings^36,37^. A half-cycle correction was applied when estimating costs and QALYs in the simulation model. The model was reviewed and validated by exploring the distributions of the simulated baseline characteristics, incidence of preventive actions, and incidence of CVD and T2D events by scenarios and risk categories from the simulation results obtained (see Supplementary File section 6 for details). The model was iteratively updated when errors were detected.

### Ethics statement

To ensure data privacy and compliance with ethical standards, this study was conducted in accordance with the Declaration of Helsinki and used only aggregated summary data derived from Nightingale Health’s analyses of Finnish THL Biobank data^38^, including the national FINRISK Studies of 2002, 2007, and 2012. The original FINRISK studies received appropriate ethical approvals from the Coordinating Ethics Committee of the Hospital District of Helsinki and Uusimaa (diary numbers: FINRISK 2002: HUS 558/E5/2001, FINRISK 2007: HUS 229/E5/2006, FINRISK 2012: HUS 238/13/03/00/2011) and and THL Biobank from the National Committee on Medical Research Ethics (Tukija) (diary number 1/2014)^39^. All participants provided informed consent, including permission for secondary use of their data. THL Biobank data was accessed through projects BB2017_98 and THLBB2023_15, under approvals granted by the relevant authorities. All statistical analyses involving individual-level data were conducted by Nightingale Health.

The research team had access only to aggregated summary data obtained from published sources and result tables generated by Nightingale Health’s data analytics team; no individual level data were accessed. Therefore, this simulation study did not require additional institutional review board approval, because it did not involve identifiable human participants or new data collection.

### Model overview

The microsimulation approach was selected as the baseline risk calculations demanded several baseline characteristics to be modelled, and the history of morbidity affects individuals’ transition probabilities between health states (see Supplementary File section 1.3 and Figure S1 for details), quality of life effects, and the resulted costs. With it, it was possible to simulate development of the individual characteristics, morbidity development, and the effects of applied preventive measures, and use the same individual as a control (i.e., an identical twin without intervention effects). The development of conditions and individual characteristics, survival, and initiation of primary prevention medication was modelled for every individual in one-year cycles until death or 100 years of age. The probabilities for developing a condition and survival varied according to the characteristics and health history of the individual. Assumptions of no regression to previous disease states were applied.

### Target population

The baseline characteristics of the target population were simulated, and the underlying “true” risks for CMD were estimated using data (n = 10,288) from the nationally representative FINRISK Studies conducted in 2002, 2007, and 2012^39^, available through the THL Biobank. Definitions of the characteristics used to define the baseline population are described in detail in Supplementary File in Table S1. Synthetic baseline characteristics of the target population were created by sampling individuals from a multinormal distribution.

### Baseline risks for T2D and CVD

For each individual, clinical risk scores, FINRISK for CVD and FINDRISC for T2D, as well as their MRS for CVD and T2D as the sum of risk factors weighted by their coefficients from the corresponding regression models were computed.

During an annual cycle of the simulation model, the onset of T2D and initial CVD event (CHD or stroke) was assessed (Table S2). Parametric survival models were fitted separately for each target disease utilizing data from the national FINRISK Studies of 2002, 2007, and 2012. The survival function was defined based on the time from recruitment to the first disease event, death, ten years from the recruitment, or to the end of follow-up, whichever came first. Age at recruitment, sex, clinical risk score (FINRISK for CHD and stroke (see Tables S3–S5) or FINDRISC for T2D (see Tables S3 and S4, Figure S2)) and metabolomic risk score (CVD specific for CHD and stroke and T2D specific for T2D) were used as model predictors. Log-logistic distribution was selected to model disease-free survival of T2D and stroke and the Weibull distribution to model disease-free survival of CHD over time based on visual inspections and information criteria (AIC, BIC) values (see Supplementary File section 2.1.3; Tables S6–S7 and Figures S3–S4 for details).

### Risk of T2D-related microvascular complications

A parametric survival regression modelling approach was applied to longitudinal real-world electronic health record data to model the annual risk of T2D-related microvascular complications in newly diagnosed disease-free T2D patients (n=7,859)^40^. The gamma distribution was selected for the modelling of a complication-free survival time based on visual examination and information criteria (AIC, BIC) values (see Table S8 for details).

### Risk of death

The national life tables conditional on age and sex (adjusted for CVD- and T2D-related deaths) were used to characterize the annual risk of all-cause death (i.e., a background mortality) in the model (see Supplementary File section 2.3 and Table S17 for details). In addition, deaths in the modelled health states were adjusted with previously published hazard ratios to consider the increased risk of death in those health states^41–44^. Deaths in incident MI, angina or stroke were modelled with reported Finnish case fatalities (Table S18)^45^.

### Modelling risk factor development over time

In the simulation model, the age of modelled individuals was updated in every annual cycle as it is an important risk factor in the modelled conditions. Since multiple other factors, such as comorbidities and use of primary prevention, also affect the risks, several annual adjustments were made (Supplementary File Tables S9–S16, Figure S5) to update the variables and the probabilities of the conditions in every cycle in the model. Furthermore, the impacts of prevalent comorbidities (e.g., an increased risk of a CVD in individuals with T2D) were considered in the model by updating the risk of CVD events in every cycle (see Supplementary File section 2.2.2 for details).

### Intervention effects

The modelled preventive actions targeted individuals at moderate and high risk of type T2D and CVD (see Supplementary File section 3; Figures S6–S8 and Tables S19–S20). In the base-case analysis, preventive actions were defined according to Finnish Current Care Guidelines^46–48^ and focused on modifying key risk factors, such as high blood pressure, hyperlipidemia, and smoking (Table S21). In a base-case analysis, preventive actions for individuals in moderate and high-risk groups were assumed to be identical in both study arms.

As part of the scenario analysis, enhanced preventive actions were modelled, incorporating stricter thresholds for high blood pressure and hyperlipidemia, along with additional interventions targeting overweight, obesity, and unhealthy diet (see Supplementary File section 3.3 for details). These enhanced actions were assumed to be implemented in both study arms. The effects of these interventions on T2D and CVD risks were estimated based on published studies^49,50^. To ensure comparability, all preventive actions in the enhanced study arms were standardized, with no differences in the interventions or medical therapies provided between study arms. However, moderate-risk individuals in the MRS-based health check arm were allocated to a nurse health check to obtain Healthy weight coaching eHealth programme.

### Health economic data inputs

Cost of nurse health check, common laboratory tests connected to it, and physician’s consultation were estimated with the costs related to them in public and private occupational health care weighted with the proportion of the health checks within the provider (Supplementary File Table S22)^25^. The costs were estimated with rates of occupational health care within private providers and health promotion visits in public health care. The cost of MRS testing was obtained from the technology provider and set at €30 per individual.

The medication costs for primary prevention were defined as the costs of antihypertensives and/or statins described in detail in Supplementary File section 4.2, Table S23. Additionally, for primary prevention, the cost of a control visit to physician’s and nurse’s office for the first year, and every other year for the following years was added for modest approach, according to the Finnish standard health care costs^51^. The control visit costs were added only once even if both antihypertensives and cholesterol medication were used.

For smoking cessation intervention among smokers, 12-week varenicline treatment was modelled as a cost without the value tax together with one visit to a physician.

The costs of T2D were derived from previously published studies^52,53^, with the exception of medication costs, which were obtained from the Dispensations reimbursed under the National Health Insurance (NHI) scheme register^54^. These costs encompass the additional healthcare expenses and productivity losses attributable to T2D and its microvascular complications (Supplementary File section 4.3, Table S24).

The costs of initial CVD events were defined based on a previous Finnish real-world study^55^, which provides the cost estimates of the hospital treatment of patients with T2D for an acute phase and the year following an initial CVD event (Table S25). Costs of case fatality due to CVD events was determined based on the official national unit cost tariff list^56^. In addition, the costs of rehabilitation and productivity losses were added to the cost of the treatment for non-fatal CVD events (Tables S26–S28). The probability of receiving a rehabilitation after a CVD event was defined based on the proportion of patients with a CVD event in rehabilitation observed in a previous Finnish national registry study^57,58^. Productivity losses associated with a CVD event were estimated by applying a human capital approach^59^. The productivity losses were added to the total costs of individuals under 65 years with the CVD event. Costs of a CVD event following the first event were estimated as annual average costs caused by medication and estimated need for health care visits based on Finnish Current Care Guidelines (Table S29)^47,56,60^. The cost of medication after CHD event was estimated from Dispensations reimbursed under the NHI scheme register^54^. The costs of one visit to physician’s and nurse’s office per year was added to medication costs^51^. The visit costs also included the possible laboratory expenses, such as the costs of cholesterol tests. As there are notable differences in the treatment and secondary prevention of intracranial haemorrhage (ICH) and ischemic stroke (IS), the weighted average based on the proportion of ICH and IS survivors, according to event case numbers and their case fatalities was used to evaluate the costs after stroke event^23,37,51^. For simplicity, a modest estimation of one visit to the nurse’s office per year was added as follow-up cost to both ICH and IS. For antithrombotic medication costs after IS, the annual cost of the combination of acetyl salicylic acid and dipyridamole was added, as it is the first option for secondary prevention in the Current Care Guidelines in Finland^47^. The yearly costs of antihypertensives and statins were retrieved from Dispensations reimbursed under the NHI scheme register^54^. As the incidence of subsequent CVD events was not considered, the costs of the treatment of subsequent CVD events were not included in the model. CVD costs are described in more detail in Supplementary File section 4.4.

### Quality of life data inputs

Utility estimates applied to estimate the number of QALYs were obtained from previously published studies (Supplementary File Table S30). Applied utility values were based on an EQ-5D-3L instrument. Baseline utilities and disutility due to diabetes and its complications were modelled as in the previous diabetes-model^52,53^. Baseline utilities varied by age and sex. Disutility associated with diabetes-related complications was estimated as a prevalence-weighted mean of disutility of each complication. Disutility associated with a CVD event during the initial event year was obtained from a previous study^61^. Disutility values after the first year were modelled with disutility values from Finnish survey study with 8,028 participants where the disutility connected to prevalent chronic conditions was studied^62^.

### Statistical analysis

Three scenario analyses were conducted to investigate the impact of several alternative assumptions on the workload effects and cost-effectiveness results:

1. Base-case analysis where standard health check including preventive actions according to the Finnish Current Care Guidelines was replaced with MRS-based health check;
2. Scenario analysis 2 where standard health check including preventive actions according to the Finnish Current Care Guidelines was replaced with MRS-based health check with enhanced preventive actions;
3. Scenario analysis 3 where standard health check with enhanced preventive actions was compared with MRS-based health check with enhanced preventive actions.

To study the robustness of the simulation results, a probabilistic sensitivity analysis (PSA) using a Monte Carlo microsimulation with 10,000 iterations of 120,000 individuals was applied to evaluate the impact of simultaneous variation in model parameters on the cost-effectiveness results. The results of the PSA were presented using cost-effectiveness acceptability curve (CEAC) calculated from the net monetary benefit statistic across a range of willingness-to-pay (WTP) thresholds^63^. The CEAC describes the probability that the “true” ICER estimate will be below the selected WTP.

### Sensitivity analyses

Two sensitivity analyses were carried out. First, attendance rate equivalent to that of standard health checks (i.e., 76.0% over five years) was utilized also for MRS-based health checks. Second, a nurse visit of 20 minutes was added to the moderate- and high-risk individuals in the MRS-based health checks to reflect the current practise used in Finnish healthcare.

### Statistics and Reproducibility

The analyses were performed using R (version 4.4.3)^64^. Detailed descriptions of the statistical methods are provided above and in the Supplementary File. Reproducibility was assessed by rerunning the microsimulation model to ensure that the findings were consistent.

## Results

### Cohort characteristics

The generated synthetic cohort of 256,372 individuals aged 50–54 years resembles of the original cohort of 10,288 participants aged 30–64 years of the national FINRISK Studies of 2002, 2007, and 2012 (Table 1).

**Table 1.** Characteristics of the original cohort and the generated synthetic data. Data are n (%) or mean (SD). Abbreviations: CVD, cardiovascular disease; HDL, high-density lipoprotein; MRS, metabolomic risk score; T2D, type 2 diabetes.

|  | Original cohort (30–64-year-olds) (n=10,288) | Original cohort (50–54-year-olds) (n=1,342) | Synthetic data (50–54-year-olds) (n=256,372) |
|---|---|---|---|
| Sex (men) | 44.8% | 45.8% | 48.4% |
| Age, years | 47.0 (10.0) | 52.0 (1.1) | 51.9 (1.4) |
| Current smokers | 23.6% | 23.5% | 23.8% |
| Body mass index (kg/m<sup>2</sup>) | 26.6 (4.3) | 27.1 (4.3) | 27.1 (4.3) |
| Height (cm) | 169.4 (9.3) | 168.7 (9.4) | 169.0 (9.2) |
| Weight (kg) | 76.7 (15.2) | 77.4 (15.2) | 77.6 (14.8) |
| Waist circumference (cm) | 89.8 (13.0) | 91.7 (12.8) | 91.8 (12.7) |
| Family history of myocardial infarction | 22.1% | 25.4% | 25.3% |
| Family history of stroke | 11.7% | 15.8% | 15.8% |
| Family history of diabetes | 24.4% | 26.8% | 26.6% |
| Systolic blood pressure (mmHg) | 130.2 (17.2) | 133.3 (17.4) | 133.9 (16.2) |
| Total cholesterol (mmol/l) | 5.5 (1.0) | 5.7 (1.0) | 5.6 (1.0) |
| HDL cholesterol (mmol/l) | 1.5 (0.4) | 1.5 (0.4) | 1.5 (0.4) |
| Antihypertensives | 11.6% | 14.9% | 14.9% |
| Lipid-lowering medication | 5.2% | 5.6% | 5.6% |
| Conventionally estimated risk for CVD and T2D (FINRISK and FINDRISC) |  |  |  |
| Low | 66.2% | 59.8% | 50.9% |
| Moderate | 18.9% | 27.1% | 33.6% |
| High | 14.9% | 13.0% | 15.5% |
| MRS-based CVD and T2D risk |  |  |  |
| Low | 69.4% | 61.3% | 58.7% |
| Moderate | 19.8% | 27.3% | 28.8% |
| High | 10.8% | 11.3% | 12.5% |

### Workload effects of MRS-based health checks

In the base-case analysis, the simulation results showed that the use of MRS-based health checks to assess the elevated risk of CMD leads to potential savings in health professionals’ workload compared with the current practices (Table 2). As shown in Table 2, the MRS-based health checks resulted in a significant reduction in nurse and physician work time compared to standard health checks in the target population.

**Table 2.** Time required for risk identification and associated FTE gains. Time required to identify individuals at risk in a 50–54-year-old population (n=256,372), comparing standard and MRS-based health checks. Values represent total hours for nurses, physicians, and laboratory work over five years, and corresponding full-time equivalent (FTE) working years gained. One FTE equals 1,600 hours. In the standard health check, risk identification includes laboratory testing (10 min) and a nurse visit (1 h), with a subsequent physician visit (30 min) for high-risk individuals. In the MRS-based health check, risk identification is based on laboratory testing (10 min), followed by a physician visit (30 min) for high-risk individuals. Abbreviations: FTE, full-time equivalent; MRS, metabolomic risk score.

| Arm | Nurse (h) | Physician (h) | Laboratory (h) | Gained nurse FTE years | Gained physician FTE years |
|---|---|---|---|---|---|
| Standard health check | 193,140 | 14,934 | 32,190 |  |  |
| MRS-based health check | 0 | 12,091 | 32,474 |  |  |
| Difference between MRS-based and standard health check | -193,140 | -2,843 | 284 | 120.7 | 1.8 |

### Long-term cost-effectiveness of MRS-based health checks

The results of the base-case analysis can be found in Table 3. With the current standard of care, the target cohort of 256,372 individuals projected a total of 3,999,095 QALYs over lifetime. If individuals would undergo MRS testing instead of conventional health checks, 1,316 QALYs could be expected to be gained (i.e., additional years in full health). Within this timeframe, the estimated total costs of standard and MRS-based health checks were €3,962 million and €3,935 million, respectively. Thus, the MRS-based health check is a likely cost-effective (i.e., more effective and less costly).

**Table 3.** Incremental costs and QALYs of MRS-based health check versus standard health check. . Savings, gained QALYs and ICER/QALY in the target population of 50–54-year-old individuals in labour force (n=256,372). Abbreviations: ICER, incremental cost-effectiveness ratio; MRS, metabolomic risk score; NA, not applicable; QALY, quality-adjusted life year.

|  | Life years per individual | QALYs, per individual | Total costs per individual, € | Difference in QALYs per individual | Gained QALYs in the target population | Difference in costs per individual, € | Savings in the target population, € | ICER/QALY, € |
|---|---|---|---|---|---|---|---|---|
| Base-case analysis |  |  |  |  |  |  |  |  |
| Standard health check | 31.8 | 15.60 | 15,453 |  |  |  |  |  |
| MRS-based health check | 31.8 | 15.60 | 15,350 | 0.01 | 1,316 | -104 | -26,567,084 | Dominant |
| Scenario analysis 2 |  |  |  |  |  |  |  |  |
| Standard health check | 31.8 | 15.60 | 15,453 |  |  |  |  |  |
| MRS-based health check | 31.9 | 15.64 | 14,448 | 0.04 | 10,813 | -1,006 | -257,803,391 | Dominant |
| Scenario analysis 3 |  |  |  |  |  |  |  |  |
| Standard health check | 31.9 | 15.65 | 14,590 |  |  |  |  |  |
| MRS-based health check | 31.9 | 15.64 | 14,448 | -0.01 | -1,672 | -142 | -36,354,488 | NA |

If MRS-based health checks would include enhanced preventive actions against CMD (scenario analysis 2), 10,813 QALYs could be expected to be gained, and the estimated total costs of MRS-based health checks would be €3,636 million resulting in savings of €258 million. If both standard health checks and MRS-based health checks would include enhanced preventive actions against CMD (scenario analysis 3), the estimated total costs of standard and MRS-based health checks were €3,740 million and €3,704 million, respectively. Although €36 million would be saved if individuals would undergo MRS testing instead of conventional health checks, 1,672 QALYs could be expected to be lost due to differences in baseline risk stratification between compared approaches (see Table 1 for details).

Estimated CEACs indicated that the MRS-based health check was likely to be cost-effective compared with standard health checks across all evaluated scenarios (Fig. 2, Supplementary Data). In the base-case, at a WTP threshold of €0 per QALY gained, the probability of cost-effectiveness was 74.5%, suggesting that the intervention was cost-saving in most simulations. However, substantial uncertainty remained, as reflected by the fact that approximately one quarter of simulations did not indicate cost savings. At a WTP threshold of €50,000 per QALY gained, the probability of cost-effectiveness was 68.1%. These results suggest that the MRS-based health check may provide economic benefits, although uncertainty around the magnitude and consistency of these benefits should be acknowledged.

**Fig. 2.**
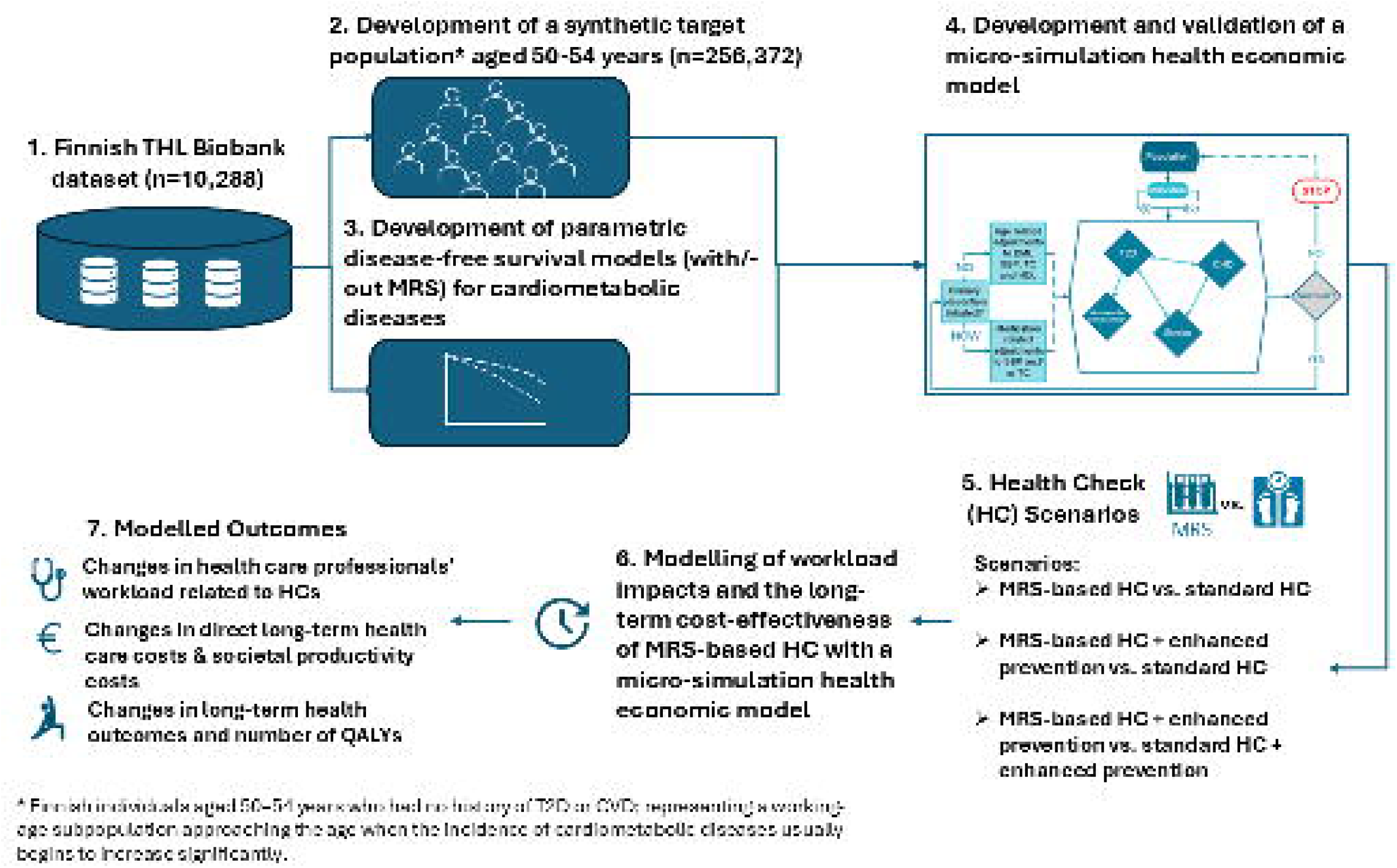
Cost-Effectiveness Acceptability Curve (CEAC). The CEAC shows the probability of the MRS-based health check to be cost-effective across selected Willingness-to-Pay thresholds as compared to standard health check.

### Sensitivity analyses

In the first sensitivity analysis assuming equivalent attendance rate in both the standard and MRS-based health checks (i.e., 76.0% over five years), the results resembled those of the primary analyses (Supplementary File Tables S31–S32). In the second sensitivity analysis, adding a nurse visit of 20 minutes to the moderate- and high-risk individuals in the MRS-based health check reduced the nurse work time savings and the savings in the target population when compared with primary analyses (Supplementary File Tables S33–S34). Altogether, interpretations of the results from sensitivity analyses remained the same with MRS-based health check being likely cost-effective option in the base-case analyses and scenario analyses 2.

## Discussion

Our study reveals that the implementation of an MRS-based health check for identifying CMD (i.e., T2D and CVD) among the working-age target population in Finland is likely a cost-saving approach compared with a standard health check. This is primarily due to the reduced expenses and quicker access due to the MRS testing approach, which offers also similar sensitivity compared with existing risk assessment tools. This leads to an enhanced early detection of disease risks, potentially decelerating the incidence rate of the targeted diseases. Furthermore, the implementation of the MRS-based health check strategy would result in substantial reductions in the health care professionals’ labour time required to identify individuals at risk for CMD. This is a critical consideration, especially in situations where there is a shortage of healthcare professionals, as is currently the case in Finland, as well as in many other countries. In addition, our results demonstrate that early detection through MRS-based health checks, combined with enhanced preventive measures, represents the optimal approach, maximizing both health benefits and cost savings. Our results also suggest that even if preventive actions were fully optimised within the current standard of care, the MRS-based health check strategy could still yield cost savings, with only minimal negative health impacts resulting from differences in risk classification. Moreover, it is important to note that a fully optimised current practice is an unlikely scenario given the ongoing cost containment pressures in Finland’s healthcare system. In contrast, the MRS-based health check strategy - when combined with optimized preventive interventions - could generate significant savings and health benefits, primarily due to reduced costs associated with risk assessment.

To our knowledge, this is the first study evaluating the workforce impacts and long-term cost-effectiveness of MRS-based health check of chronic preventable diseases. Therefore, the possibilities for comparing our results with other studies are currently limited. Previously published health economic evidence consistently suggests that overall early detection and preventive interventions of CMDs are predominantly cost-effective strategies supporting the findings of our study^65–68^. As our results show, ensuring that all conditions are managed according to current clinical guidelines once detected would further increase the health benefits without substantially increasing the costs. The benefits of early detection and management for each high-risk condition vary depending on several factors, including the proportions currently undiagnosed, the proportions currently poorly managed, the costs and effectiveness of the key management interventions, and the increased risk for CVD and other complications that each condition confers.

Although the analyses were conducted in a Finnish population, conventional risk assessment tools originating from Finland, such as the FINDRISC score, have been externally validated and widely implemented across diverse European and non-European populations^30,69–73^, supporting the broader applicability of metabolic risk assessment and its associations with cardiometabolic disease outcomes. Nevertheless, we acknowledge that risk prediction performance and absolute risk estimates may vary according to population structure, ancestry, socioeconomic factors, and healthcare system characteristics. The transferability of findings related to workload and cost-effectiveness is likely more limited. Differences in primary care organization, staffing structures, consultation lengths, and unit costs may substantially affect efficiency estimates. Importantly, the relatively low unit costs and structured nature of the Finnish publicly funded primary care system^74,75^ may lead to conservative estimates of potential cost savings compared with other European settings with higher personnel costs.

Our study has strengths and weaknesses. The key strength of our study is its basis in a single modelling framework and the use of nationally representative longitudinal biobank datasets to determine the baseline characteristics of the target population, as well as to model the risks of target diseases conditionally on applied baseline risk scores. However, there are also several limitations. First, there are some inaccuracies remaining in the modelling of the MRS risk of individuals and modelled parametric disease-free survival times (i.e. the ability of MRS to recognize the ones in true risk seems to be a bit weaker in our model than reported in previous studies^15,16^). This makes the results presented weaker than they ought to be. Second, there are some limitations regarding the applied assumptions in our study. For example, for simplicity, we assumed a one-time risk assessment, even though disease risks may increase over time, for example, due to the aging of the target population. Implementing repeated risk assessments, such as every five years, could enhance the resource efficiency of the MRS-based approach, as fewer resources would be required for each subsequent assessment cycle. A limitation of the analysis is that no prospective head-to-head comparison between the MRS and conventional screening approaches was available. Consequently, differences in downstream outcomes relied on the associations estimated from the available cohort data rather than direct comparative evidence. In the present study, we modeled a single closed cohort of individuals, whereas, in reality, new individuals enter the target age group each year. Therefore, the scalability advantages of the MRS-based approach could be even greater when applied at the population level over time. Since the incidence of subsequent CVD events was not accounted for in the present study, the costs associated with treating these events were not included in the model. This may lead to an underestimation of long-term CVD costs, as recent real-world evidence suggests that, for example, a T2D diagnosis combined with a prior CVD event increases the risk of recurrent CVD events and the associated healthcare costs^60^. Finally, our analysis captures only a portion of the potential value of the MRS-based approach, as the same MRS testing also provides risk estimates for numerous other preventable diseases^15,16^ beyond the scope of this study. For instance, MRS-based risk assessments are already used in clinical practice in both Finnish occupational health services and public health care for unemployed individuals^28,29^ to identify elevated risks for several different preventable diseases, thereby supporting the early detection and prevention of multimorbidity. This is particularly important given that individuals with multimorbidity represent one of the fastest growing and most costly patient groups for health systems^76^.

Our study highlights some equity issues within the Finnish healthcare system. Funded primarily through taxation, healthcare services are provided by wellbeing services counties, which are responsible for both primary and specialized care. Public social and health services are supplemented by private providers, which currently account for over a quarter of all social and health services in Finland. Under Finnish law, employers are required to provide preventive occupational healthcare for all employees. These services, predominantly delivered by private occupational health care providers, play a significant role in disease prevention and health promotion among the working-age population. However, access to these services remains limited for unemployed individuals, leading to disparities in preventive healthcare. Therefore, the broader implementation of scalable low cost first-line risk assessments could enhance access to preventive services while addressing equity concerns in a resource-efficient manner.

While our study provides novel insights into the workforce impacts and long-term cost-effectiveness of the MRS-based health check in improving the detection and management of high-risk individuals, further research is needed to assess the real-world outcomes and feasibility of MRS-based health checks, including laboratory capacity requirements, integration into care workflows, training needs, and clinician and patient acceptability, as well as monitor potential unintended consequences such as risk assessment-related anxiety in prospective implementation studies. These further developments would also offer valuable insights for other healthcare systems by identifying key enablers and barriers to the adoption of the MRS-based health check approach, ultimately supporting the transition toward more proactive and preventive healthcare systems.

## Acknowledgments

The authors would like to acknowledge the help from Sini Kerminen, PhD, Nightingale Health PIc, for conducting the analyses based on the THL Biobank data and for providing the resulting aggregated outputs to the research team.

## Data availability

All data used in the analyses are provided in the Supplementary File, and the data underlying Figure 1 are available in the Supplementary Data. Most of the input data were derived from previously published studies. In addition, the study utilized data from the THL Biobank, including the FINRISK 2002, 2007 and 2012 studies. Due to legal and ethical restrictions, individual-level biobank data cannot be shared publicly. Researchers may apply for access to these data through the THL Biobank and the relevant data access authorities in accordance with Finnish legislation and applicable data permit requirements.

## Code availability

The code used in this study is not publicly available due to intellectual property and related restrictions. The analytical methods are described in sufficient detail to enable replication of the main findings. Reasonable requests regarding code availability may be directed to Piia Lavikainen, with an anticipated response time of up to four weeks.

## Authors’ contributions

PL: formal analysis, methodology, supervision, validation, visualisation, writing – original draft. LH-S: methodology, resources, software, validation, visualisation, writing – original draft. AVL: formal analysis, methodology, software, visualisation, writing – original draft. KJ: methodology, resources, software, writing – review & editing. JH: project administration, methodology, writing – original draft. TL: supervision, writing – review & editing. JM: conceptualisation, funding acquisition, methodology, resources, supervision, writing – original draft.

## Supplementary File

**Supplementary File. Health Care Workload Impacts and Cost-Effectiveness of a Metabolomic Risk Score-based Health Check for Cardiometabolic Disease Prevention in Finland.**

**Supplementary Data. Source data of Figure 2**.

