## Supplementary File for "Health Care Workload Impacts and Cost-Effectiveness of a Metabolomic Risk Score-based Health Check for Cardiometabolic Disease Prevention in Finland"

### 1 Microsimulation model

#### 1.1 Background

A holistic and diversely applicable lifetime microsimulation model for Finnish setup was created with which the cost-effectiveness of preventative measures against cardiometabolic diseases (cardiovascular disease, CVD, and type 2 diabetes, T2D) can be evaluated. The model was created to assess holistic effects of preventative measures against these common chronic non-communicable conditions through e.g., medical and lifestyle interventions.

Microsimulation was selected as a method for two main reasons acknowledged by the International Society for Pharmacoeconomics and Outcomes Research (ISPOR) and the Society for Medical Decision Making (SMDM)[1]. Microsimulation is to be used typically when the history of comorbidities affects the probabilities of contracting conditions, which is the case also with the selected chronic diseases. Another matter affecting the choice of method was the quantity of background characteristics affecting the risks of diseases.

The health economic microsimulation model is a flexible computational framework designed to simulate individual units and their probabilistic progression over time. It is structured as a collection of interconnected submodels, each representing a distinct process (such as T2D, cardiovascular disease etc.) within the system. These submodels operate sequentially – in a random order, with outputs influencing subsequent stages of the simulation. The structure allows for the addition or removal of submodels based on specific research needs while maintaining necessary dependencies between components.

#### 1.2 Population

The population of the model covers all Finnish 50–54-year-old individuals, with annually developing characteristics simulated according to the information from a Finnish national cohort study. Depending on the need for the analysis, a subgroup, e.g., individuals in a specific age-range, could be selected for analysis. To model intervention and control arms, each individual in the model is cloned, and these clones are used to represent the participants with (intervention participant, $i_i$) and without (control, $i_c$) the intervention effect. The cloning is done for every intervention arm when needed. The incidence, costs, and effects on quality-of-life of initial CVD events, T2D and its complications and the excess mortality caused by the diseases are used as outcome measures.

#### 1.3 Model structure

The generalized model structure is presented in **Figure S1**. Synthetic individuals forming the baseline population were fed to the model one-by-one. Before entering the model, the individual was cloned, and the risk reduction introduced by the intervention was issued to one of the clones ($i_i$). The individuals then journeyed through the model in one-year cycles until death or age of 100.

During an annual cycle, the onset of T2D, T2D complications, and initial CVD event (coronary heart disease (CHD) or stroke) is evaluated according to the risks calculated with the individual baseline characteristics and the metabolomic risk score (MRS) based on a metabolic biomarker profiling. The survival of individuals is assessed annually, and the assessment is based on age- and sex-related general population mortality adjusted with effects of the comorbidities developed during the time in the model. In case of incident CHD or stroke event, case fatality is added to mortality rate.

In case of survival, the development of individual characteristics due to ageing is also modelled, and the initiation of primary prevention for CVD, defined as antihypertensive and/or lipid-lowering medication, is tested using threshold values for risk factors based on clinical guidelines. If primary prevention is initiated the characteristics targeted, i.e., blood pressure and/or cholesterol, are adjusted accordingly in the model.

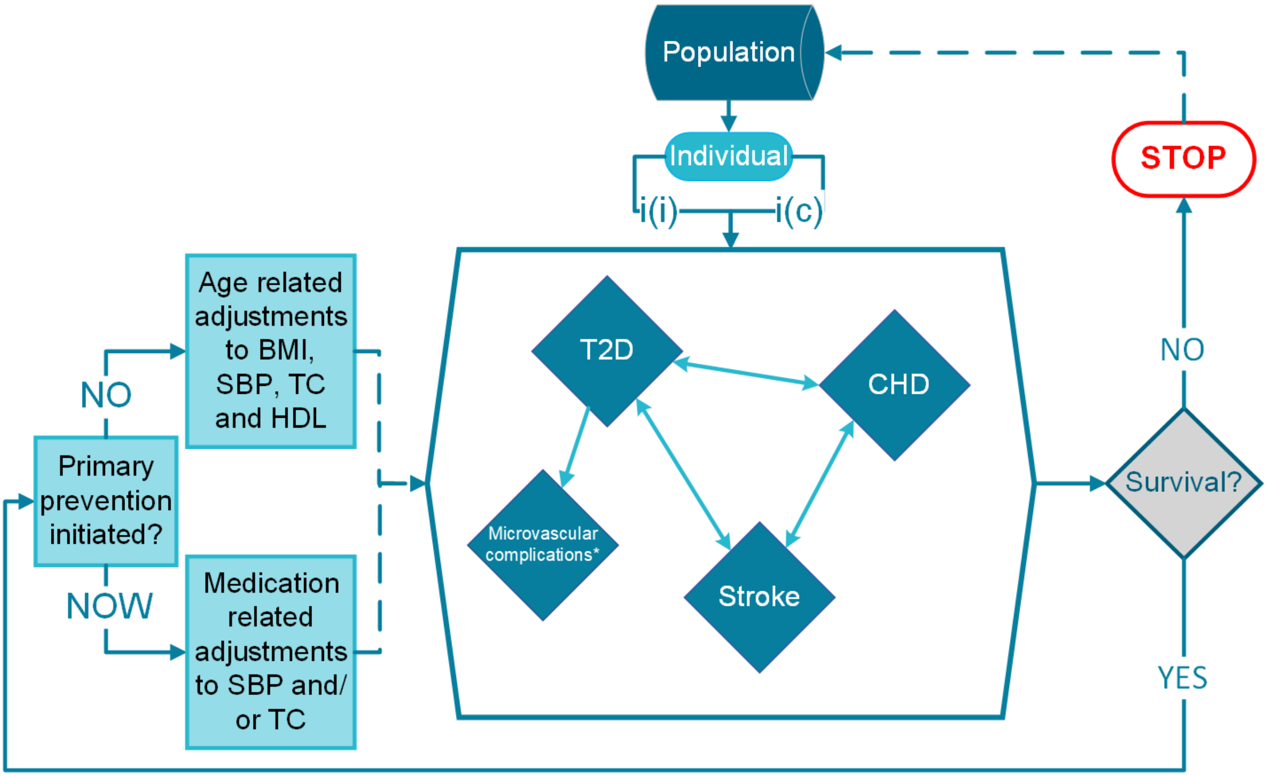

* Microvascular complications include diabetic neuropathy, foot complications, retinopathy and other eye complications, and diabetic nephropathy.
BMI, body mass index; CHD, coronary heart disease; HDL, high density lipoprotein; i(c), individual (control); i(i), individual (intervention); SBP, systolic blood pressure; TC, total cholesterol, T2D, type 2 diabetes

**Figure S1.** Structure of the microsimulation health economic model for a primary prevention of cardiometabolic diseases.

#### 2 Population

For the present study, the model was populated with individuals aged 50–54 years. The total population in this age group in Finland in 2023 was 315,799 individuals (159,989 males and 155,810 females)[2]. 89% of the population aged 50–54 years are included in labour force[3] and 9.29% of the population has prevalent CVD or T2D[4]. When assuming that 4.7% and 24.2% of those with prevalent CHD and stroke, respectively, are outside of labour force[5,6], the proportion with CVD or T2D in the labour force is 8.78%. Thus, the final size of the target population is 256,372 individuals.

The baseline characteristics of the target population were simulated based on real-world data from the national FINRISK Studies of 2002, 2007, and 2012[7], representative of Finnish population (**Table S1**). Samples from these studies were first filtered to a subset of individuals aged 30–64 years, who had complete information for age at recruitment, sex, body mass index, waist circumference, systolic blood pressure, cholesterol measurements, smoking status, family history of myocardial infarction and stroke, usage of blood pressure medication, and metabolomic variables). Individuals with underlying diabetes, CHD or stroke events at the time of recruitment were excluded, resulting in a sample size of 10,288 individuals.

For each individual, clinical risk scores, FINRISK[8] for CVD and FINDRISC[9] for T2D, as well as their metabolomic risk scores (MRS) for CVD and T2D (Nightingale Health Plc risk scores currently utilized in Finnish health care settings, including occupational health care and public health services for the unemployed[10,11]; methodology described in *Nightingale Health Biobank Collaborative Group, 2024*[12]) were simulated based on the distributions that were calculated as the sum of risk factors weighted by their coefficients from the corresponding regression models from the FINRISK Studies.

Simulated target populations were created by sampling individuals from a multinormal distribution. Means and covariance matrices were estimated separately for males and females using data described above. Continuous variables (age at recruitment, body mass index weight, height, systolic blood pressure, cholesterol measurements) were log-transformed before estimating the parameters. The dichotomous variables were treated as continuous variables assuming that the variables follow a normal distribution. For simplicity, an assumption of linearity of the relations of the modelled characteristics was made.

**Table S1.** Definitions of the characteristics modelled for the baseline population.

| Variable | Variable type | Definition | Origin of the data |
|---|---|---|---|
| **Body mass index (kg/m$^2$)** | continuous | Measured in study | FINRISK 2002, 2007 & 2012 |
| **Weight (kg)** | continuous | Measured in study | FINRISK 2002, 2007 & 2012 |
| **Height (cm)** | continuous | Measured in study | FINRISK 2002, 2007 & 2012 |
| **Waist circumference (cm)** | continuous | Measured in study | FINRISK 2002, 2007 & 2012 |
| **Systolic blood pressure (mmHg)** | continuous | Measured in study. Mean of the second and the third measurement. | FINRISK 2002, 2007 & 2012 |
| **Total cholesterol (mmol/l)** | continuous | Measured in study | FINRISK 2002, 2007 & 2012 |
| **High-density lipoprotein cholesterol (mmol/l)** | continuous | Measured in study | FINRISK 2002, 2007 & 2012 |
| **Smoking** | dichotomous | Self-reported. Yes, if current smoker. No, if non-smoking, quit smoking over 1/2 years ago, quit smoking less than 1/2 years ago. | FINRISK 2002, 2007 & 2012 |
| **Family history of diabetes** | dichotomous | Self-reported. Parental diabetes. | FINRISK 2002, 2007 & 2012 |
| **Family history of myocardial infarction** | dichotomous | Self-reported. MI in father before the age of 60 years or in mother before the age of 65 years. | FINRISK 2002, 2007 & 2012 |
| **Family history of stroke** | dichotomous | Self-reported. Parental stroke before the age of 75 years. | FINRISK 2002, 2007 & 2012 |
| **Use of cholesterol medication** | dichotomous | Self-reported. Current use of cholesterol lowering medication. | FINRISK 2002, 2007 & 2012 |
| **Use of blood pressure medication** | dichotomous | Self-reported. Use of antihypertensive medication during the last 7 days. | FINRISK 2002, 2007 & 2012 |

#### 2.1 Baseline risks for T2D and cardiovascular disease

##### 2.1.1 Definitions

The conditions modelled in the present study are defined in **Table S2**. CHD includes acute coronary syndrome (myocardial infarction and, unstable angina), and deaths because of ischaemic heart disease, cardiac arrest, and unwitnessed death. Stroke includes ischaemic stroke and intracranial haemorrhage. The definition of CVD includes CHD and stroke.

For individuals' disease status, we used disease outcomes pre-defined by THL biobank based on national hospital and cause-of-death registries (detailed description in *Julkunen et al., 2023*)[13]. In summary, CHD cases were defined as ICD-10 codes I20, I21, I22 for main diagnosis, I21 or I22 as secondary diagnosis, coronary artery bypass grafting or angioplasty, or ICD-10 codes I20–I25, I46, R96, R98 for causes of death. Stoke cases were defined as ICD-10 codes I61, I63; not I63.6, I64 as main or secondary diagnosis or cause of death. Strokes included hemorrhagic strokes (I61), ischemic strokes (I63 excluding I63.6) and unspecified strokes (I64). T2D cases were defined as ICD-10 codes E11–E14 as main or secondary diagnosis, cause of death, or with individuals with diabetes medication. ICD-8 and ICD-9 codes corresponding to the above were also considered when defining the disease history.

**Table S2.** ICD-9 and ICD-10 codes of the modelled conditions.

| Condition | ICD-8/9 | ICD-10 | Other |
|---|---|---|---|
| **CVD** | CHD + Stroke | | |
| **CHD morbidity**<br>**CHD mortality** | 410, 4110<br>410–414, 798; not 7980A | I20.0, I21, I22<br>I20–I25, I46, R96, R98 | Coronary artery bypass grafting<br>Angioplasty |
| **Stroke** | ICD-9: 431, 4330A, 4331A, 4339A, 4340A, 4341A, 4349A, 436, ICD-8: 431 (except 43101, 43191) 433, 434, 436 | I61, I63 (not I63.6), I64 | |
| **T2D** | 250 | E11–14 | Medication (Kela) |
| **Microvascular complications** | | Eye: E11.3, H28.0, H36 (not H36.8), H40.5, H42.0, H43.1, H45.0, H54<br>Foot: I70.2, I73.9, I79.2, L97, M14.2, M14.6, N48.4<br>Neuro: E11.4, E11.5, E11.6, G59.0, G63.2, G73.0, G99.0<br>Renal: E11.2, N08.3, N18, Z49, Z94.0 | Foot: NFQ10, NFQ20, NGQ10, NGQ20, NHQ10, NHQ20, NFQ48, NGQ48, NHQ30, NHQ40, NHQ60 (NOMESCO) |

Abbreviations: CHD, coronary heart disease; CVD, cardiovascular disease; ICD, International Statistical Classification of Diseases; T2D, type 2 diabetes.

##### 2.1.2 Survival models: variables

The probability of the development of CVD or T2D in the model was estimated with parametric survival models for each condition. Age, sex, MRS CVD or T2D risk and FINRISK CHD or stroke risk or FINDRISC points were used as predictors. Survival models were implemented using data from the FINRISK studies from the years 2002, 2007, and 2012.

The original FINRISK and FINDRISC risk equations have been presented in detail elsewhere[8,9,14]. Short descriptions of data, methods and variables used in the risk equations are given in **Tables S3** and **S4**. In summary, the risk scores were originally designed as tools for health care professionals for detecting the individuals in high risk of the conditions of interest. The recognition of these high-risk individuals enables the initiation of preventive measures, such as healthy lifestyle guidance with or without medication. With preventative measures initiated early enough, there is a possibility to prevent or at least delay the onset of the condition.

**Table S3.** Overview of the risk equations used to calculate the clinical risk, i.e., the probability of contracting a chronic illness.

| Risk function | Evaluated risk | Study details | Description | Source |
|---|---|---|---|---|
| **FINRISK 2.0** | 10-year risk of CHD, stroke, and combined (CVD) | National cohort studies FINRISK 1982, 1987, 1992, 1997, 2002, and 2007 (n=39,790) with follow-up of 10 years from registries for each cohort | Logistic regression model was used to create a risk function with continuous variables. The model was created separately for men and women, and for CHD and stroke events. | [8,14] |
| **FINDRISC** | 10-year risk of drug-treated T2D | National cohort studies FINRISK 1987 (n=4746) and 1992 (n=4615) with follow-up from registries until 1997 | β-coefficients of logistic regression model were used to create risk scores for different risk factors. Easy to assess parameters were chosen to the model deliberately to ensure feasibility. Possible total scores were categorized into 4 risk groups (later updated to 5), with specified risks to report the result of the risk assessment. | [9,15] |

Abbreviations: CHD, coronary heart disease; CVD, cardiovascular disease; FINDRISK, Finnish Diabetes Risk Score; T2D, type 2 diabetes.

**Table S4.** The variables and their categories used in FINDRISC and FINRISK 2.0 risk equations.

| | FINRISK$_{CHD}$ | FINRISK$_{STROKE}$ | FINDRISC | FINDRISC points |
|---|---|---|---|---|
| **Age (years)** | continuous | continuous | 45–54<br>55–64<br>> 64[¥] | 2<br>3<br>4 |
| **Sex** | male/female | male/female | - | |
| **Body mass index (kg/m$^2$)** | - | - | > 25<br>25–30<br>> 30 | 0<br>1<br>3 |
| **Waist circumference (cm)** | - | - | Male:<br>94–102<br>≥102<br>Female:<br>80–88<br>≥ 88 | <br>3<br>4<br><br>3<br>4 |
| **Systolic blood pressure** | continuous | continuous | - | |
| **Blood pressure lowering medication[¢]** | - | - | yes/no | 2/0 |
| **Total cholesterol (mmol/L)** | continuous | - | - | |
| **High-density lipoprotein cholesterol (mmol/L)** | continuous | continuous | - | |
| **Diabetes diagnosis** | yes/no | yes/no | - | |
| **Smoking[#]** | yes/no | yes/no | - | |
| **Physical activity** | - | - | ≥4 hours/week<br><4 hours/week[%] | 0<br>2 |
| **Daily consumption of vegetables, berries, and fruits** | - | - | yes/no | 1/0 |
| **High blood glucose level at any time point** | - | - | yes/no | 5/0 |
| **Family history of the disease** | yes/no[@] | yes/no[^] | No history[¥]<br>2. degree relative[¥&]<br>1. degree relative[¥+] | 0<br>3<br>5 |

Abbreviations: CHD, coronary heart disease
[¥]category/variable added later to the risk assessment form;
[#]Smoker, regular smoking for at least a year and has smoked during the last month;
[¢]"Have you ever taken medication for high blood pressure on regular basis?";
[¤]20-30 minutes of physical activity causing sweating and breathlessness;
[%]Individuals without physically straining spare time activities and who work mainly sitting;
[&]Distant relatives, i.e. grandparents, aunts, uncles, first cousins;
[+]Close relatives, i.e. parents, siblings, children;
[@]Myocardial Infarction on a parent at age under 65 years;
[^]Stroke on a parent at age under 75 years.

FINRISK 2.0 equations were estimated based on the 10-year follow-up of 39,790 individuals[8]. CHD and stroke are predicted by smoking, systolic blood pressure, high-density lipoprotein cholesterol, diabetes, and family history, added with total cholesterol for CHD (**Table S4**). **Table S5** shows the applied coefficients of the FINRISK risk equations for CHD, stroke, and the combined CVD risk.

**Table S5.** Risk equations for the acute coronary heart disease, stroke, and cardiovascular disease (death or hospital treatment) in the next 10 years (%)[8].

| **Risk for men** | |
|---|---|
| **CHD** | $(1/(1+e^{(9.081-0.075\times age-0.579\times SM-0.320\times TC+1.082\times HDL-0.011\times SBP-0.729\times DIA-0.338\times PMI)}))\times 100$ |
| **Stroke** | $(1/(1+e^{(9.928-0.083\times age-0.369\times SM+0.329\times HDL-0.014\times SBP-0.705\times DIA-0.249\times PS)}))\times 100$ |
| **Risk for women** | |
| **CHD** | $(1/(1+e^{(11.250-0.095\times age-0.639\times SM-0.244\times TC+0.845\times HDL-0.013\times SBP-1.315\times DIA-0.421\times PMI)}))\times 100$ |
| **Stroke** | $(1/(1+e^{(9.553-0.085\times age-0.613\times SM+0.623\times HDL-0.012\times SBP-0.914\times DIA-0.023\times PS)}))\times 100$ |
| **United risk for both events** | |
| | $100\times(1-(1-\mathrm{CHDrisk})\times(1-\mathrm{STROKErisk}))$ |

Abbreviations: CHD, coronary heart disease; DIA, diabetes; HDL, high density lipoprotein; PMI, parental myocardial infarction; PS, parental stroke; SBP, systolic blood pressure; SM, smoking; TC, total cholesterol.

The FINDRISC risk score is based on baseline information from the national FINRISK study data from 1987 (n=4,746) and validated with the FINRISC study data from 1992 (n=4,615) with baseline surveys and examination[9]. Development of drug treated T2D was followed for 10 years from the Prescription Register maintained by the Social Insurance Institution of Finland. In the FINDRISC score T2D is predicted by age, BMI, waist circumference, physical activity, consumption of berries, fruits, and vegetables, use of blood pressure medication, history of high blood glucose, and family history (**Table S4**). The β-coefficients of the logistic regression model of FINDRISC were used to create scores for the risk factors[9]. The scoresheet has been updated (**Figure S2**) since the function was first published[15] with age group of over 64 years and a question about family history of diabetes.

As some of the variables needed in calculating FINDRISC (consumption of vegetables, fruits and berries, physical activity, and history of high blood glucose) were not available in our FINRISK 2002, 2007 and 2012 data, these were imputed using linear regression and variables on age, body-mass index, weight, height, waist circumference, systolic blood pressure, total cholesterol and HDL cholesterol. Regression coefficients used for imputation were based on a covariance matrix from a FinHealth 2017 study and were obtained from the National Institute of Health and Welfare (personal communication). FINRISK and FinHealth studies belong to the same series of population health studies in Finland. Information on T2D prevalence in distant relatives was not available in any of the available datasets and is missing from the baseline characteristics. Thus, the FINDRISC scores were slightly underestimated in the model.

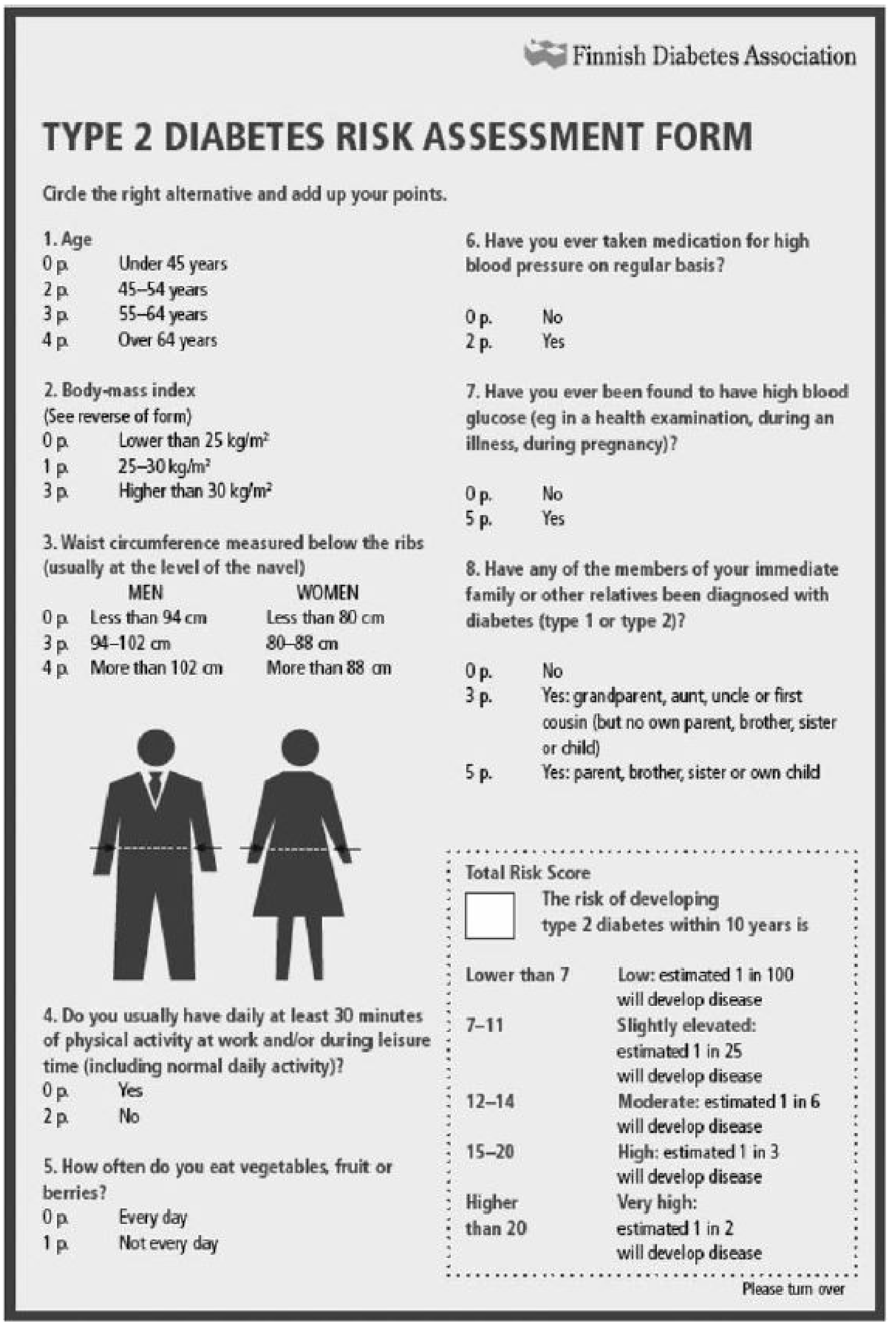

Finnish Diabetes Association

### TYPE 2 DIABETES RISK ASSESSMENT FORM

Circle the right alternative and add up your points.

**1. Age**
0 p. Under 45 years
2 p. 45–54 years
3 p. 55–64 years
4 p. Over 64 years

**2. Body-mass index**
(See reverse of form)
0 p. Lower than 25 kg/m²
1 p. 25–30 kg/m²
3 p. Higher than 30 kg/m²

**3. Waist circumference measured below the ribs (usually at the level of the navel)**

| | MEN | WOMEN |
|---|---|---|
| 0 p. | Less than 94 cm | Less than 80 cm |
| 3 p. | 94–102 cm | 80–88 cm |
| 4 p. | More than 102 cm | More than 88 cm |

**4. Do you usually have daily at least 30 minutes of physical activity at work and/or during leisure time (including normal daily activity)?**
0 p. Yes
2 p. No

**5. How often do you eat vegetables, fruit or berries?**
0 p. Every day
1 p. Not every day

**6. Have you ever taken medication for high blood pressure on regular basis?**
0 p. No
2 p. Yes

**7. Have you ever been found to have high blood glucose (eg in a health examination, during an illness, during pregnancy)?**
0 p. No
5 p. Yes

**8. Have any of the members of your immediate family or other relatives been diagnosed with diabetes (type 1 or type 2)?**
0 p. No
3 p. Yes: grandparent, aunt, uncle or first cousin (but no own parent, brother, sister or child)
5 p. Yes: parent, brother, sister or own child

Total Risk Score

The risk of developing type 2 diabetes within 10 years is

| | |
|---|---|
| Lower than 7 | Low: estimated 1 in 100 will develop disease |
| 7–11 | Slightly elevated: estimated 1 in 25 will develop disease |
| 12–14 | Moderate: estimated 1 in 6 will develop disease |
| 15–20 | High: estimated 1 in 3 will develop disease |
| Higher than 20 | Very high: estimated 1 in 2 will develop disease |

Please turn over

**Figure S2**. The scoresheet and risk classes of the FINDRISC risk calculator[15].

MRS-based CVD and T2D risks were identified with the technology provided by the Nightingale Health Plc. Nightingale Health–based MRSs are developed using high-throughput nuclear magnetic resonance (NMR) spectroscopy, which quantifies a standardized panel of circulating metabolites, including detailed lipoprotein subclasses, fatty acids, amino acids, and glycolysis-related biomarkers[12,16].

Disease-specific risk scores (e.g., for T2D, stroke, and CHD) are derived by combining metabolite concentrations with conventional risk factors (age and sex) in multivariable regression frameworks, most commonly Cox proportional hazards models for time-to-event outcomes. Variable selection is performed using a combination of prior biological knowledge and data-driven approaches, and the resulting models assign regression coefficients (β weights) to each biomarker to generate a continuous risk score.

The models are developed using large population-based cohorts, such as UK Biobank and Finnish cohort studies, and are externally validated across multiple independent cohorts, such as Estonian Biobank and Finnish THL Biobank, to ensure generalizability. Validation typically involves assessing discrimination (e.g., C-statistics) and calibration, as well as evaluating the incremental predictive value of metabolomic information beyond traditional clinical risk factors.

Overall, Nightingale MRSs provide standardized, reproducible predictors of cardiometabolic risk that complement conventional risk models by capturing underlying metabolic perturbations associated with disease development. Nightingale MRSs are already implemented in Finnish healthcare[10,11].

##### 2.1.3 Survival models: selection of the models

Using the available real-world data from the national FINRISK Studies of 2002, 2007, and 2012, parametric survival models were conducted separately for all conditions with 8 parametric survival distributions (exponential, Weibull, Gompertz, log-logistic, log-normal, generalized gamma, generalized gamma orig., and gamma) using *flexsurvreg* function of *flexsurv* package (v 2.3.2) in R. Age at recruitment, sex, clinical risk score (FINRISK for CHD and stroke or FINDRISC for T2D) and metabolomic risk score (CVD specific for CHD and stroke and T2D specific for T2D) were used as model predictors. To match FINDRISC score (often represented as risk points) to the same risk space as FINRISK and MRS that report the results as quantitative absolute disease risks, logistic regression results of the 'Full model' from Lindström & Tuomilehto, 2003[9] were utilized. Survival models to be used in the cost-effectiveness simulation were chosen based on visual examination and information criteria. Cumulative hazard plots (**Figure S3**) and 50-year survival plots (**Figure S4**) of these models were visually compared with data. Additionally, Akaike Information Criterion (AIC) and Bayesian Information Criterion (BIC) values, evaluating the fit of the model to the data, were compared with between the models (**Table S6**).

The visual examinations revealed that exponential, Gompertz, and generalized gamma original were not suitable to model any of the conditions. Other distributions had only minor differences. After closer observations of the plots and information criteria values, log-logistic distribution was selected to model T2D and stroke, and Weibull to model CHD (**Table S7**). The use of parametric disease-free survival models enabled the extrapolation of disease-free survival probabilities over true follow-up time (i.e., which is not possible with conventional non-parametric Kaplan-Meier survival curves), and thus the estimation of the annual probabilities of developing T2D or CVD over the applied lifetime horizon in the present analysis.

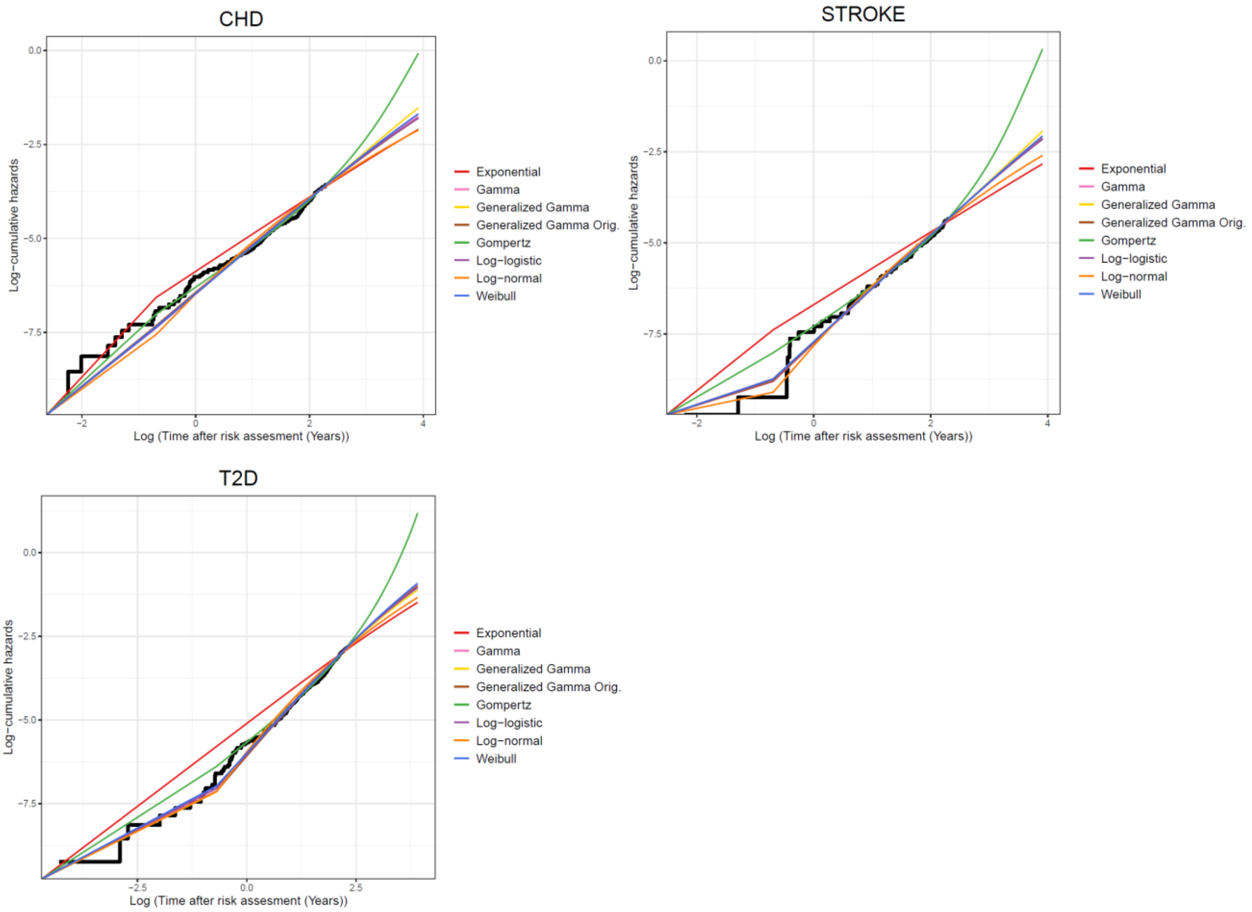

**Figure S3.** Cumulative hazard plots of the conducted parametric survival models.

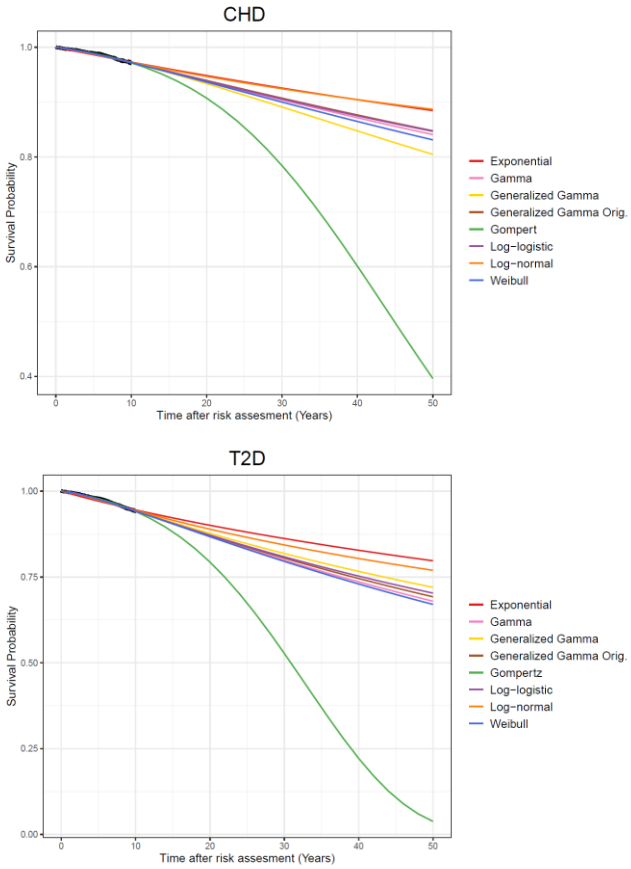

**Figure S4.** Lifetime (50-year) extrapolation of disease-free survival with the conducted parametric survival models.

**Table S6.** Information criteria values (smaller the better) of alternative survival models.

| **Distribution** | **CHD** | | **Stroke** | | **T2D** | |
|---|---|---|---|---|---|---|
| | **AIC** | **BIC** | **AIC** | **BIC** | **AIC** | **BIC** |
| **Exponential** | 2884 | 2920 | 1482 | 1518 | 5150 | 5187 |
| **Weibull** | 2872 | 2916 | 1469 | 1516 | 5092 | 5136 |
| **Gompertz** | 2868 | 2912 | 1471 | 1515 | 5087 | 5131 |
| **Log-logistic** | 2872 | 2916 | 1469 | 1512 | 5074 | 5118 |
| **Log-normal** | 2879 | 2923 | 1471 | 1514 | 5095 | 5139 |
| **Generalized gamma** | 2875 | 2923 | 1472 | 1523 | 5079 | 5130 |
| **Generalized gamma original** | 2875 | 2926 | 1472 | 1522 | 5083 | 5133 |
| **Gamma** | 2876 | 2920 | 1472 | 1516 | 5084 | 5127 |

Abbreviations: AIC, Akaike information criterion; BIC, Bayesian information criterion; CHD, coronary heart disease; T2D, type 2 diabetes.

**Table S7.** Parameters of the selected disease-free survival models.

| Parameter | Estimate | SE |
|---|---|---|
| **CHD (Weibull)** | | |
| Shape | 1.274 | 0.079 |
| Scale | 13.918 | 10.606 |
| Female sex | -0.009 | 0.157 |
| Baseline age | -0.010 | 0.010 |
| MRS-based risk | -0.091 | 0.154 |
| Clinical risk | -0.759 | 0.121 |
| **Stroke (log-logistic)** | | |
| Shape | 1.480 | 0.137 |
| Scale | 55.781 | 90.312 |
| Female sex | 0.315 | 0.183 |
| Baseline age | -0.025 | 0.018 |
| MRS-based risk | 0.237 | 0.173 |
| Clinical risk | -0.744 | 0.203 |
| **T2D (log-logistic)** | | |
| Shape | 1.496 | 0.063 |
| Scale | 4.605 | 1.140 |
| Female sex | 0.120 | 0.075 |
| Baseline age | -0.008 | 0.004 |
| MRS-based risk | -0.484 | 0.040 |
| Clinical risk | -0.326 | 0.027 |

Abbreviations: CHD, coronary heart disease; MRS, metabolomic risk score; T2D, type 2 diabetes.

##### 2.1.4 The incidence of diabetes microvascular complications

To model the incidence of T2D microvascular complications, a gamma model was created from a real-world electronic health record data from the Wellbeing Services County of North Karelia with 7,859 newly diagnosed disease-free diabetics from years 2011 and 2022 followed until the end of 2022[17] using *flexsurvreg* function of *flexsurv* package (v 2.3.2) in R. The diagnosis codes interpreted as T2D complications are presented in **Table S2** and the regression coefficients for the complication model are shown in **Table S8**. The original model calculated the risk of onset of T2D complication in days, which were changed to years for use in the model. Only the first incident complication of a person was modelled.

**Table S8.** Gamma regression coefficients for T2D microvascular complications.

| Parameter | Estimate | SE |
|---|---|---|
| **Shape** | 1.090 | 0.037 |
| **Rate** | <0.001 | <0.001 |
| **Female sex** | 0.242 | 0.067 |
| **Baseline age** | 0.027 | 0.003 |

#### 2.2 Risk and risk factor development

The survival models were used to model the risk for the selected conditions based on age, sex, MRS risk and FINRISK risk or FINDRISC points. Of these, age was updated in every cycle as it is important risk factor in the modelled conditions. Since multiple other factors, such as comorbidities and use of primary prevention, affect also the risks, a number of annual adjustments was made (**Table S9**) to update the variables and the probabilities of the conditions in every cycle in the model.

**Table S9.** Assumptions relating to the development of risk factors over time.

| Yearly adjusted variable | Direct effects | Secondary effects |
|---|---|---|
| **Age** | Directly affects the quantity of weight change, true risk of all the conditions and death. | Affects SBP, TC, and HDL change through weight change. |
| **BMI/weight** | Directly affects the change in SBP, TC, and HDL.<br>Weight loss through HWC directly affects the true risk of T2D. | Weight-loss through HWC affects the true risks of CHD and stroke through changes in SBP, TC, and HDL. |
| **SBP** | Directly affects the assessment of need for antihypertensive medication in primary prevention. | Affects the assessment of need for lipid-lowering medication in primary prevention through FINRISK 2.0. |
| **TC** | Directly affects the assessment of need for lipid-lowering medication in primary prevention. | Affects the assessment of need for lipid-lowering medication in primary prevention through FINRISK 2.0. |
| **HDL** | | Affects the assessment of need for lipid-lowering medication in primary prevention through FINRISK 2.0. |
| **FINRISK 2.0 combined CVD risk** | Directly affects the assessment of need for lipid-lowering medication in primary prevention. | |
| **Antihypertensive medication** | Directly affects the true risks of CHD and stroke. | |
| **Lipid-lowering medication** | Directly affects the true risks of CHD and stroke. | |
| **"True" risk of CHD** | Directly affects the probability of getting CHD during the cycle | |
| **"True" risk of stroke** | Directly affects the probability of getting stroke during the cycle | |
| **"True" risk of T2D** | Directly affects the probability of getting T2D during the cycle | |
| **"True" risk of death** | Directly affects the probability of dying during the cycle | |

Abbreviations: CHD, coronary heart disease; HDL, high-density lipoprotein cholesterol; HWC, healthy weight coaching; SBP, systolic blood pressure; T2D, type 2 diabetes; TC, total cholesterol.

##### 2.2.1 Modelling the trajectories of the risk factors

The change in background characteristics was modelled through change in BMI. The annual change in BMI was modelled according to *Pajunen et al., 2010*[18] who examined the annual change in weight based on FINRISK surveys from 1972 to 2007. The magnitude of change in weight until 69 years of age was dependent on sex, age and current BMI group of the individual as shown in **Table S10**. Normal distribution was assumed. After turning 70 years, no change was assumed.

**Table S10.** Annual change in weight (kg) according to sex, age and BMI[18].

| **Age group** | **BMI<25** | | **25≤BMI<29.9** | | **BMI≥30** | |
|---|---|---|---|---|---|---|
| **Men** | Change in kg | 95% CI | Change in kg | 95% CI | Change in kg | 95% CI |
| **25–39** | 0.48 | 0.41–0.54 | 0.40 | 0.31–0.49 | 0.36 | 0.09–0.64 |
| **40–49** | 0.30 | 0.21–0.38 | 0.26 | 0.18–0.35 | 0.37 | 0.11–0.63 |
| **50–69** | 0.30 | 0.16–0.44 | 0.11 | 0.01–0.21 | -0.15 | -0.40–0.10 |
| **Women** | | | | | | |
| **25–39** | 0.47 | 0.41–0.52 | 0.52 | 0.38–0.65 | 0.41 | 0.20–0.61 |
| **40–49** | 0.42 | 0.35–0.49 | 0.35 | 0.27–0.43 | 0.23 | 0.05–0.41 |
| **50–69** | 0.16 | 0.05–0.27 | 0.05 | -0.08–0.19 | 0.08 | -0.11–0.27 |

Abbreviations: BMI, body mass index; CI, confidence interval

Changes in BMI over time were modelled to affect systolic blood pressure, total cholesterol, and high-density lipoprotein cholesterol levels. The effect of change in BMI to the waist circumference was not modelled. The changes in other variables were modelled through changes in BMI according to pooled analysis by[19] of two lifestyle intervention trials executed in Europe (**Table S11**).

**Table S11.** Pooled effects of 1 kg weight loss to other variables[19] applied in the model.

| **Variable** | **Change in variable/kg loss in weight** | **95 % Confidence Interval** |
|---|---|---|
| **Systolic blood pressure (mmHg)** | -0.4 | -0.45 to -0.32 |
| **Total cholesterol (mmol/L)** | -0.02 | -0.02 to -0.01 |
| **High-density lipoprotein cholesterol (mmol/L)** | 0.003 | -0.0006 to 0.006 |

##### 2.2.2 Effects of comorbidities on the disease risk

Comorbidities were modelled to affect the probability to contract CVD and T2D. Thus, the baseline probability of a condition was adjusted accordingly as other conditions developed. The adjustments and their sources are presented in **Table S12**.

**Table S12.** Adjustments made on disease probability according to prevalent morbidity.

| Condition contracted | Condition with adjusted probability | Risk modification (95 % CI) | Source |
|---|---|---|---|
| **Diabetes** | CHD event<br>men<br>women | <br>RR 1.98<br>RR 3.57 | Calculated with FINRISK 2.0 [8] |
| | Stroke<br>men<br>women | <br>RR 1.97<br>RR 2.44 | Calculated with FINRISK 2.0 [8] |
| **CHD event** | Stroke | First year: HR 5.3<br>After first year: HR 1.5 | Weighted averages from [20] |
| **Stroke** | CHD-event | HR 2.0 | [21] |

Abbreviations: CHD, coronary heart disease; CI, confidence interval; CVD, cardiovascular disease; HR, hazard ratio; RR, relative risk.

For simplicity, the incidence of secondary cardiovascular event, similar to the first one (CHD or stroke), was not explicitly modelled in the present model. However, CVD event of different type was modelled as there is no protective effect with CHD on stroke or vice versa. On the contrary, the probability of the events is increased by incidence of the other[20,21]. After CHD-event, the probability of stroke was accommodated with adjusted rate ratios for ischaemic stroke and ICH after myocardial infarction from Danish population[20]. The HRs gained through weighted average will probably slightly overestimate the number of strokes as in our model the CHD events include unstable angina. The risk was modelled separately for the first year after the MI and from then on. Post-stroke CHD-event risk was accommodated with unadjusted, combined HR for MI and unstable angina pectoris from Canadian population-based cohort study by *Sposato et al., 2020*[21].

##### 2.2.3 Effects of medication

In the present model, primary prevention was defined as treatment for hypertension and/or hyperlipidaemia initiated before an actual CVD event. The Current Care Guidelines were used to determine who should be treated for hypertension and/or hyperlipidaemia according to the variables included in the model[22,23]. For example, in the model, the need for antihypertensive medication was possible to define through SBP only, even though also diastolic blood pressure is used in real world.

The effects of the medication were modelled based on results from previous clinical studies. For each risk factor (i.e. systolic blood pressure and total cholesterol) the risk reduction is based on the change that can be achieved with the medication. The initiation of each medication and the effects of them in the model are presented in detail below.

###### 2.2.3.1 Antihypertensive medication

Initiation of antihypertensive medication was assumed for every individual experiencing CHD or stroke regardless of the level of systolic blood pressure[24,25]. In primary prevention with antihypertensive medication the initiation limit was set to 140 mmHg[22]. As already mentioned above, diastolic blood pressure had to be ignored here because it was not modelled. According to the Current Care Guidelines, a lifestyle guidance is given, and lifestyle change experimented first if systolic blood pressure exceeds 140 mmHg but is under 160 mmHg, and medication initiated only if this is not enough to lower the blood pressure. For simplicity this lifestyle change was not modelled. However, as all individuals in need of the medication do not initiate it due to, e.g., ignorance of the need, or negative attitudes towards medication use, the probability of initiating the treatment was calculated using the proportion of people on medication among those with prescription for antihypertensive medication, or systolic blood pressure over 140 mmHg according to the FinHealth 2017 study[26] (**Table S13**). The process of initiation of antihypertensive medication is presented in **Figure S5a**.

According to *Ahola et al., 2016*[27], the average number of antihypertensive medication in use per patient in Finland is 1.8 (in 2011). Thus, the effect of medication was averaged assuming 1.8 drugs was automatically used for every individual starting the treatment. This will underestimate the treatment of the most severe cases of hypertension patients but following the findings of the FinHealth study with only 42.5% of men and 41.8% of women achieving the usual target of under 140/90 mmHg even if treated[26], this was considered a compromise we could accept.

**Table S13.** Use of antihypertensive medication of Finnish people with prescribed antihypertensive medication or blood pressure over 140/90 mmHg by sexes[26].

| | Men | Women |
|---|---|---|
| **Use of antihypertensive medication during past 7 days (%)** | 37.8 | 44.4 |

The effect of antihypertensive medication on SBP in individuals initiating the treatment, was calculated using an equation[28], where the effect depends on the level of blood pressure before initiation of drug treatment. The average decrease in systolic blood pressure that the first antihypertensive inflicts was defined with

$$\text{Decrease in SBP} = 9.1 + 0.10\times(\text{pSBP} - 154)\ \text{mmHg},$$

where pSBP is the pretreatment systolic blood pressure. For the following drug, the additional effect is calculated with the same equation, but with the pSBP replaced with the gained SBP after initiating the first drug and the gained decrease multiplied with 0.8 to account for the total effect of 1.8 drugs. After initiation of antihypertensive, no annual changes in blood pressure were implemented. The risk reductions were implemented with values from a published study[29] and are presented in **Table S14**.

**Table S14.** Risk reduction achieved with antihypertensive medication.

| Condition | Unit of change | HR | 95% CI | Reference |
|---|---|---|---|---|
| **CHD** | -5 mmHg | 0.92 | 0.89–0.95 | [29] |
| **Stroke** | -5 mmHg | 0.87 | 0.84–0.91 | [29] |

Abbreviations: CHD, coronary heart disease; CI, confidence interval; HR, hazard ratio.

###### *2.2.3.2 Lipid-lowering medication*

Initiation of lipid-lowering medication was assumed for every individual experiencing CHD or stroke regardless of the level of total cholesterol or other lipids[24]. The Current Care Guidelines for ischaemic stroke primarily recommend lifestyle guidance and change, and medication initiation only if considered necessary[25]. Again, for simplicity, this lifestyle change was not modelled. Also, as stroke in our model also includes ICH, this procedure is not quite adequate, but it was still considered satisfactory as the types of strokes were not modelled separately.

a.

b.

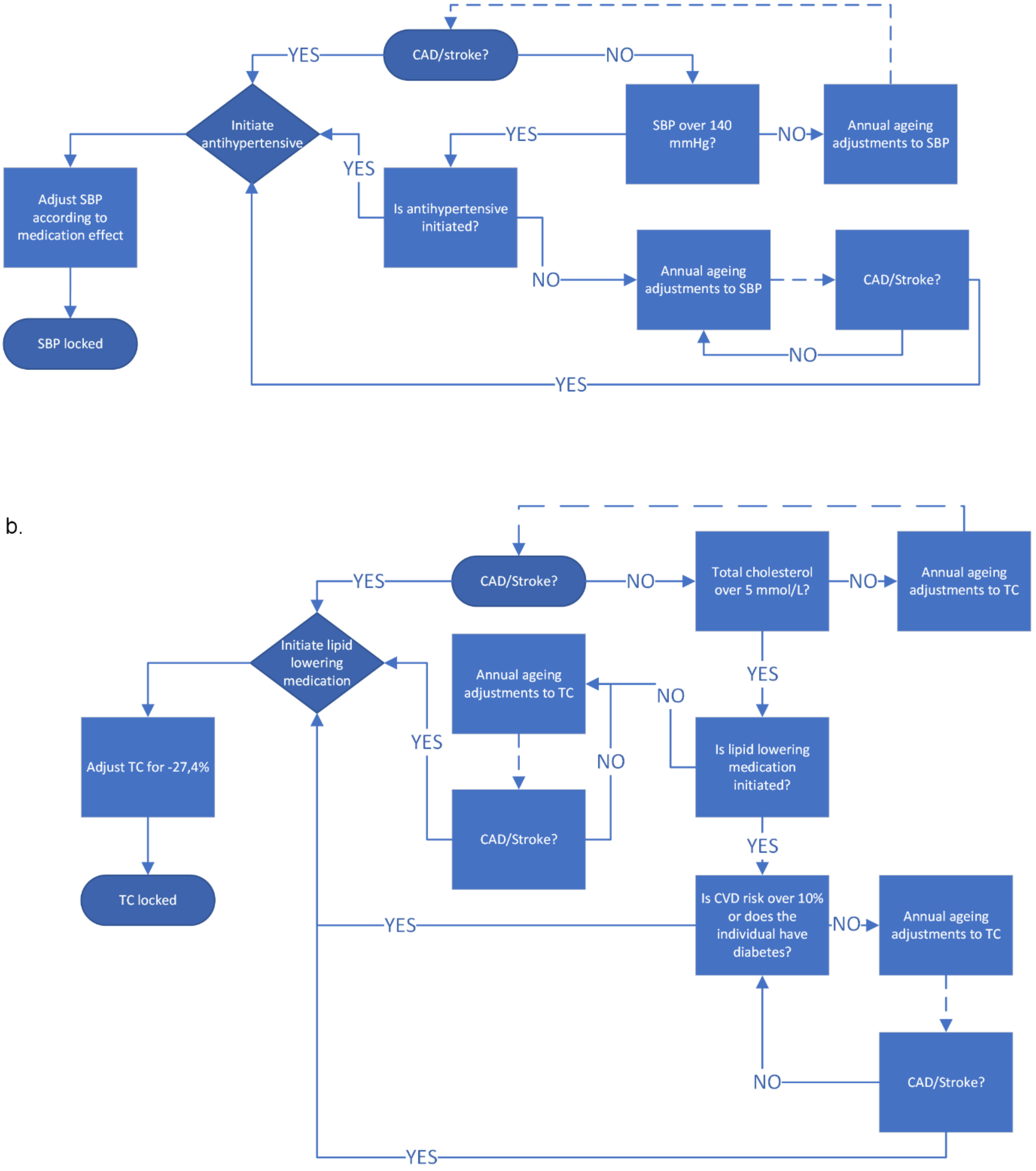

CAD, coronary artery disease; CVD, cardiovascular disease; SBP, systolic blood pressure; TC, total cholesterol

Same cycle

Cycle change

**Figure S5.** Applied processes of initiating antihypertensive (a.) and lipid lowering medication (b.) in the present model.

According to the Finnish Current Care Guidelines, the drug treatment for hyperlipidaemia should be initiated in primary prevention if cholesterol level is abnormal and the total risk for CVD, calculated with the FINRISK 2.0 equation presented in **Table S5**, is over 10% for the next 10 years[23]. This is automatically considered to be true if the individual has diabetes. However, lipid-lowering medication is not initiated for everyone who satisfies these conditions as the abnormal cholesterol levels may not be discovered, and not everyone in need for the medication uses it. This was modelled by using the proportion of medication users in potential 50–59 year-old population for the medication use, i.e., population with cholesterol over 5.0 mmol/L, or cholesterol lowered to normal level with medication, as the probability for initiating the lipid lowering medication (**Table S15**)[26].

**Table S15.** Details on cholesterol levels, lipid-lowering medication use, and the probability to use lipid lowering medication if individuals' natural cholesterol is over 5.0 mmol/L in 50–59 year-old population[26].

| | Men | Women |
|---|---|---|
| **Abnormal cholesterol levels (% of the entire population)** | 64.3 | 76.5 |
| **On medication (% of the entire population)** | 19.1 | 9.9 |
| **Reached objective with medication (LDL<3mmol/L) (% of those on lipid-lowering medication)** | 78.8 | 72.9 |
| **Target population for lipid-lowering medication (those with abnormal cholesterol level or cholesterol levels lowered to normal level with medication) (% of the entire population)** | 79.4 | 83.7 |
| **On medication of the target population (%)** | 24.1 | 11.8 |

Abbreviations: LDL, low-density lipoprotein cholesterol.

The process of initiation of lipid-lowering medication in our model is presented in **Figure S5b**. In summary, CHD or stroke automatically initiated the medication. Otherwise, if individual's total cholesterol reached 5.0 mmol/L, the individual was deemed to initiate or not to initiate with lipid-lowering medication. If deemed initiating, the combined CVD risk of that individual was evaluated annually. If it was over 10%, or the person had T2D, the treatment for hyperlipidaemia was initiated. If the individual is deemed not to initiate the treatment, the treatment is not initiated at any time point, except after CVD-event as part of normal care.

The risk reduction after initiating lipid-lowering medication was calculated with estimated effect of the medication on LDL using TC as a proxy. Thus, initially the effect of the medication on total cholesterol was modelled. The full effect of statins was modelled as 27.4% reduction in total cholesterol values. The effect was calculated as weighted average of the mean effects of commonly used doses of different statins in trials[30] (**Table S16**). The doses match quite well to the ones used in Finland in the beginning of 2000s[31]. Effect of two different doses of rosuvastatin, 5 mg and 10 mg, was given, the smaller was used to avoid overestimating the effect. The distribution of different active substances of statins in use in Finland calculated from prescription register of statistical database Kelasto maintained by Social Insurance Institution of Finland was used as weight[32]. The change of active substance by a user could not be taken into consideration, and thus, the number of users is an overestimation. However, according to previous research this will have only a minimalistic effect on the proportions[33]. After adjustments of TC levels due to statins, no change in TC was assumed to take place.

**Table S16.** Effect size and proportion of use in Finland of different statins. The numbers were used to calculate the weighted average effect of statins[30,32].

| Statin | Effect size (% change from baseline) | Proportion of users in Finland in 2021 (%) |
|---|---|---|
| **Atorvastatin** | 27 | 41.0 |
| **Fluvastatin** | 17 | 1.2 |
| **Lovastatin** | 23 | 0.3 |
| **Pravastatin** | 21 | 1.6 |
| **Rosuvastatin 5 mg** | 30 | 29.6 |
| **Simvastatin** | 26 | 26.3 |

It has been shown previously that 1.0 mml/L reduction in LDL cholesterol is associated with a 1.2 mmol/L reduction in TC[34]. This association was used to calculate the reduction in LDL from reduction in TC. HR 0.78 per 1 mmol/L reduction in LDL was used to model the risk reduction after initiating lipid-lowering medication[35,36]. This HR has been used also in the LIFE-CVD model[37].

###### *2.2.3.3 Blood glucose lowering medication*

The objective of drug therapy in T2D is to treat the blood glucose level to avoid symptoms and complications of the condition[38]. The effects of diabetes medication were included in the incidence of complications estimated based on a previous Finnish study[17]. As the trajectory of blood glucose level was not modelled, there was no need to model the effect of medication on it.

#### 2.3 Mortality

The background mortality of general population was obtained from the Finnish-specific lifetables and defined as age and gender-specific all-cause mortality (except for T2D- and CVD-related mortality) from the year 2022 and applied in the developed model[39,40]. Background mortality was adjusted with disease-specific mortality when required (**Table S17**).

**Table S17.** Additional mortality related to the modelled conditions.

| Condition | HR (95% CI) | Source |
|---|---|---|
| **T2D** | | |
| **Women** | 2.47 (2.23–2.72) | [41] |
| **Men** | 1.93 (1.79–2.07) | [41] |
| **T2D with complications** | 2.36 (1.70–3.29) | [42] |
| **ACS** | 1.3 | Systematic review[43] |
| **Stroke** | 1.74 (1.06–2.85) | [44] |

Abbreviations: ACS, acute coronary syndrome; CI, confidence interval; HR, hazard ratio; T2D, type 2 diabetes.

The risk of death of individuals with T2D was adjusted with sex-related HR from a previous study[41]. Furthermore, the risk of death of individuals with T2D with complications was adjusted additionally with OR from a previous study[42]. After surviving MI, mortality was adjusted with a coefficient from a review studying the long term (>10 years) mortality in MI survivors (≥one year)[43]. For stroke survivors, the mortality was adjusted with a coefficient from a population based NHANES study with 10,025 participants in the USA[44].

Finnish-specific estimates for case-fatality associated with angina, MI, and stroke were considered in the model and used to account for a higher risk of death in the year when the event occurred (**Table S18**). The information was retrieved from Performance, Effectiveness and Cost of Treatment episodes (PERFECT) reports which include data from, e.g., the Finnish Hospital Discharge Register (THL) and Cause of Death Register (Statistics Finland) and is described in detail elsewhere[45–47]. The data includes patients in yearly cohorts that have been treated in hospital for initial acute coronary syndrome (ACS, ICD-10: I20.0, I21, I22) or stroke (I60, I61, I63, I64) or died because of CHD (I20-I25, I46, R96, R98, R99) or stroke (I60, I61, I63 (excl. 163.6), I64). Patients in long-term care in an institution before the event are excluded from the data. In the PERFECT reports initial cases are defined as the patient not being treated in hospital for the same condition for 365 days before the hospital admission or death.

Case-fatality associated with ACS was applied to the cohort developing ACS and stroke-related mortality was applied to the cohort developing stroke event. Case-fatality was defined as death that occurs within 30 days of the ACS event and for stroke events as death that occurs within 180 days of the event.

**Table S18.** Case fatality of initial cardiovascular events.

| CVD event | Death probability | Death within | Source |
|---|---|---|---|
| **Case fatality ACS** | 0.073 | 30 days | [48] (reported year 2019) |
| **Case fatality stroke** | 0.151 | 180 days | [48] (reported year 2018) |

Abbreviations: ACS, acute coronary syndrome (myocardial infarction, unstable angina); CVD, cardiovascular disease.

#### 3 Intervention and control arms

To compare the MRS-based health check (mHC) against the standard health check (sHC), two alternative study arms were modelled. Outlines of both arms are presented in **Figure S6**. In the sHC arm, individuals attended conventional health checks, including a laboratory visit and a nurse visit. During the nurse visit, moderate- and high-risk individuals were identified according to the Finnish Current Care Guidelines using FINDRISC and FINRISK risk scores as part of the standard health check process in Finland. In mHC arm, MRS testing was used instead of conventional health checks. Participants were then classified into three classes (high, moderate and low) depending on their CVD and T2D risk (the highest risk class of the two is used) assessed with clinical risk tests in sHC arm and MRS blood testing in mHC arm. In a base-case analysis, preventive actions for individuals in moderate and high-risk groups were assumed to be identical in both study arms and follow the national Current Care Guidelines. In scenario analyses, enhanced preventive actions were implemented.

The aims of the simulations were to model

1) the impact of MRS-based risk identification instead of the conventional health check,
2) the added value of MRS-based risk identification with enhanced preventive actions instead of the conventional health check, and
3) the added value of conventional and MRS-based health checks combined with enhanced preventive actions.

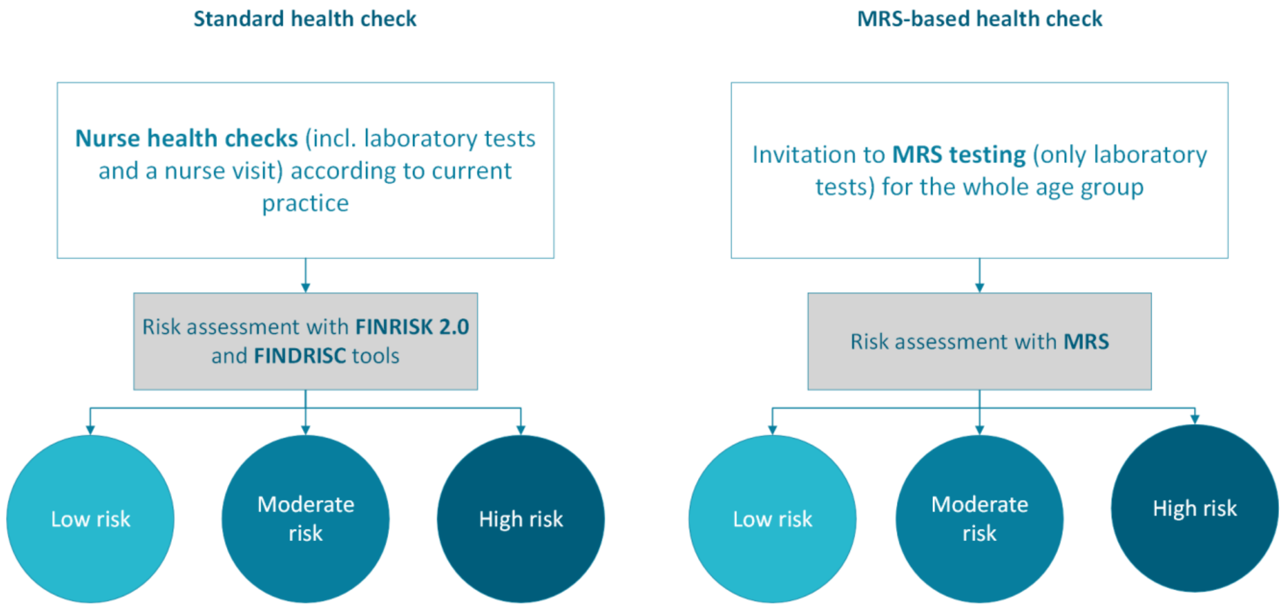

**Figure S6.** Risk identification in standard and metabolomic risk score (MRS)-based health checks.

##### 3.1 Participation

In the sHC group individuals attended nurse health checks according to current practice in Finland: independently through occupational health care or through public health care services for the unemployed, or as invited by occupational health care service provider. In 2021, there were 398,009 nurse health checks in occupational health care, excluding the health checks due to exposure in job assignments[49]. This is 15.6% of the employed work force[50]. 8.6% of the unemployed work force had a free health check in the public health care[51]. The unemployment rate of the age group was 5.3% in 2023[52]. Thus, the overall attendance rate was calculated as a weighted average: 15.6% × (100% − 5.3%) + 8.6% × 5.3% =15.2%. This 15.2% annual attendance rate was used as a base-case assumption of for sHC during the first 5 years of the model. In mHC arm, the whole population aged 50–54 years was invited to MRS risk testing to laboratory in the first year. The attendance rate for this small and low contribution testing was assumed to be similar as with the standard health check (total of 76.0% (5*15.2%) of target population for 5 years) but within the first year of the model due to saved worktime resources.

#### 3.2 Classification

The risk assessment in the sHC group was done with the traditional clinical risk calculators (i.e., FINRISK and FINDRISC risk equations). In real life, these assessments are not done in every health check. There is no information on the extent of their use available. In the present study, an assumption of use in every nurse health check was made. The FINRISK risk classes for CHD and stroke were defined according to Finnish Current Care Guidelines with 10-year probability of combined CVD risk (**Table S19**). FINDRISC risk classes were defined as in the scoresheet (**Figure S2**) by combining low and slightly elevated into low-risk group, and high and very high into high-risk group.

**Table S19.** CVD and T2D risk groups used in health care practice according to Current Care Guidelines.

| | Low | Moderate | High |
|---|---|---|---|
| FINRISKI CVD-risk (CHD+stroke) | <5% | 5–10% | >10% |
| FINDRISC points (T2D) | 0–11 | 12–14 | 14–26 |

Abbreviations: CHD, coronary heart disease; CVD, cardio-vascular disease; T2D, type 2 diabetes.

In the mHC arm, MRS testing was used to assess the risk of CVD and T2D. MRS testing takes place in laboratory through blood sampling. MRS risk classes were defined so, that the proportion of individuals in all three risk classes are approximately the same in both sHC and mHC arms (**Table S20**).

**Table S20.** Limits used in the risk classification of the mHC arm.

| | Low | Moderate | High |
|---|---|---|---|
| CVD-risk (CHD+stroke) | <4.25% | 4.25–7.75% | ≥7.75% |
| T2D risk | <3.46% | 3.46–6.28% | ≥6.28% |

Abbreviations: CHD, coronary heart disease; CVD, cardio-vascular disease; T2D, type 2 diabetes.

It was assumed that mHC was delivered through a digital healthcare application that automatically provided personalized risk information and prompted individuals identified as high risk to book a physician appointment. Under this scenario, no dedicated nurse appointment was required for risk assessment or communication of results because risk identification was based on a blood sample and the test results were returned electronically. It was assumed that administrative tasks were integrated into existing digital systems and therefore required minimal additional healthcare personnel time compared with the sHC.

Agreement between conventional and MRS-based classifications was moderate (Cohen's $\kappa = 0.428$) in the synthetic data. While 66.8% of individuals were classified into the same risk category by both approaches, 33.2% were reclassified, demonstrating that the MRS-based strategy does not simply reproduce conventional risk classification. Furthermore, metabolomics-based risk scores have previously been shown to improve discrimination and risk reclassification beyond conventional clinical risk scores for both CVD and T2D, with reported improvements in C-index and net reclassification indices[53,54].

#### 3.3 Preventive actions

Preventive actions were, as mentioned, were similar in the sHC and mHC strategies (**Figure S7**). Preventive actions targeted individuals at moderate and high risk of type T2D and/or CVD. In the base-case analysis, these actions were defined according to Finnish Current Care Guidelines[22,25,38] and focused on modifying key risk factors, such as high blood pressure, hyperlipidemia, and smoking. All individuals in each of the risk groups were assumed to undergo the intervention if the prespecified risk limits were met (full adherence). For smokers, a smoking cessation intervention was modelled despite of the identified risk class. It was assumed to contain 12-week rehabilitation treatment with varenicline. A modest success rate of 15% was assumed for the smoking cessation based on review observation on abstinence rate of varenicline at 6 months (21.8%) and added with an assumption that some relapses would happen thereafter[55].

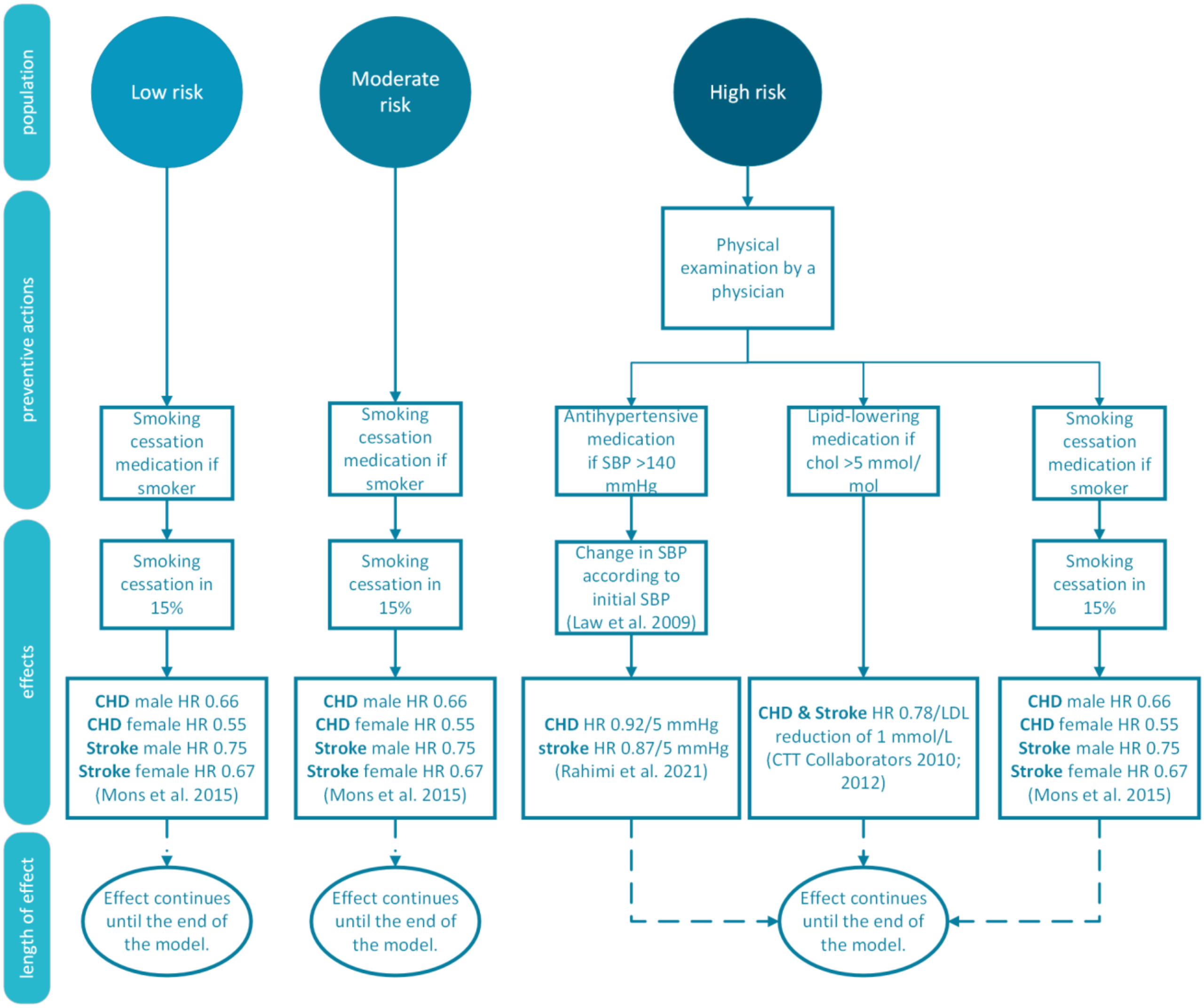

**Figure S7.** Preventive actions in the base-case analysis. Abbreviations: CHD, coronary heart disease; SBP, systolic blood pressure.

In other scenario analyses, enhanced preventive actions were implemented (**Figure S8**). For moderate risk group, lifestyle intervention through the Healthy weight coaching (HWC) eHealth programme was initiated. HWC is a lifestyle intervention with 12-month interactive web-based program which handles broad spectrum of health behaviours such as diet, physical activity, psychological factors, and coping with stress[56]. It includes, for example, training sessions and personal coach. In the mHC group HWC was preceded with a nurse health check.

For high-risk group, preventive actions included the same HWC eHealth programme for all and smoking cessation intervention for smokers but also a health check with physician to assess the need for primary prevention. Lipid-lowering medication was initiated for all in the high-risk group and antihypertensive medication was initiated, with no extra limitations, if SBP was over 130 mmHg.

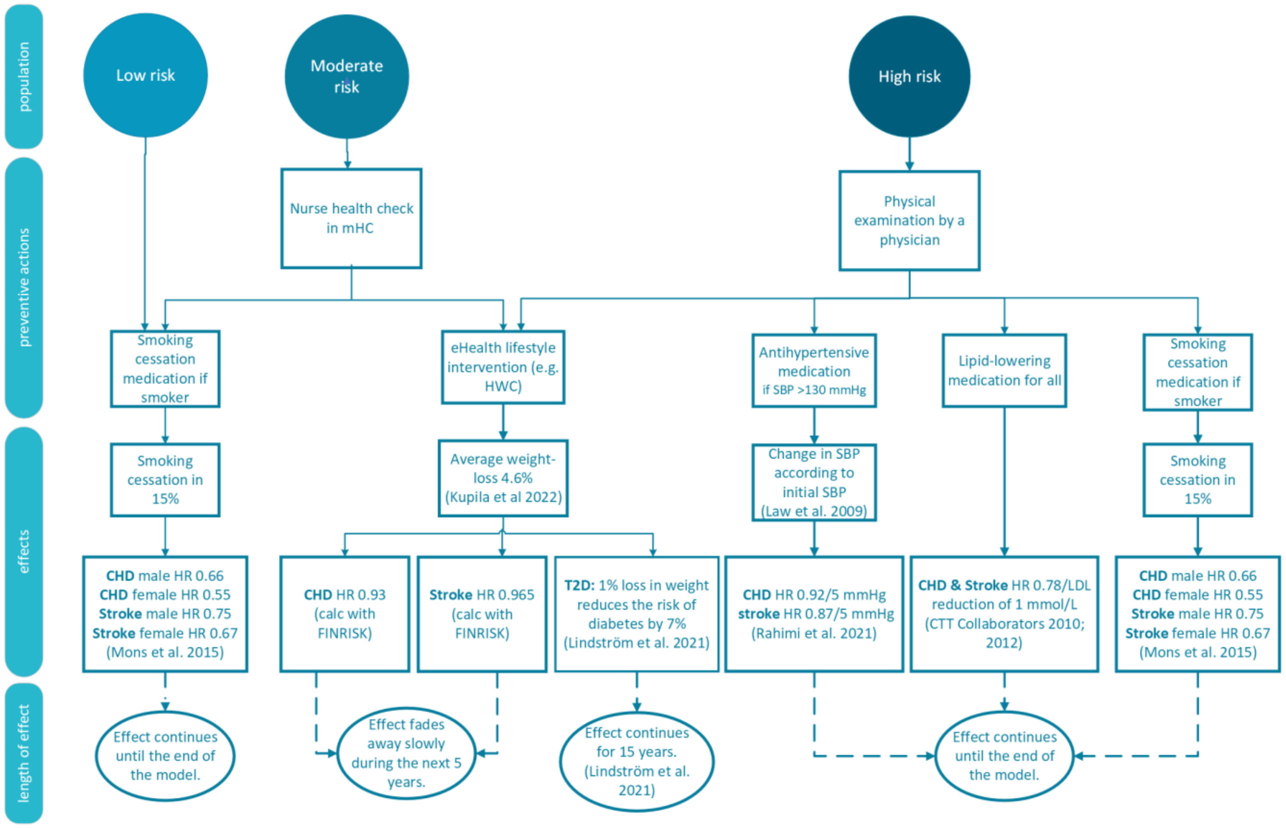

**Figure S8**. Enhanced preventive actions in scenario analyses. Abbreviations: CHD, coronary heart disease; HWC, Healthy weight coaching; SBP, systolic blood pressure.

##### 3.3.1 Risk reduction

The efficacy of the preventive actions was introduced in the model through changes in BMI and cardiometabolic risk factors (i.e., SBP, HDL cholesterol, total cholesterol) and risk reductions from the published studies applied directly to the risk. The main driver of efficacy of the HWC eHealth intervention was the reduction in weight and BMI. The mean change of weight after 12 months of HWC was 4.6% (SE 0.5%)[56]. Changes in BMI influence the risk of T2D directly, and the risk of CVD through changes the BMI reduction causes in other variables, i.e., SBP, total cholesterol, and HDL. To account for the effects of the weight loss on the risk of T2D, the risk of T2D was modified by relative risk estimates, which were derived from a paper by *Lindström et.al., 2021*[57]. They concluded that on grounds of the Diabetes Prevention Study (DPS) analysis, 1% loss in weight reduces the risk of diabetes by 7%. To model the decrease in CHD and stroke, the effect of change in BMI on CVD risk factors included in FINRISK 2.0 was calculated. Risk ratios of the new calculated risks for CHD (0.93) and stroke (0.965) separately and the old ones was then used to model the risk reduction.

For smoking cessation a risk reduction was modelled according to meta-analysis on impact of smoking and smoking cessation on CVD events[58] (**Table S21**). As the effect of smoking was already included to baseline risk modelling through FINRISK 2.0, only the risk reduction following smoking cessation compared to those continuing to smoke was added to the model separately for the sexes and for CHD and stroke. For antihypertensive and lipid-lowering medication, the risk reduction was modelled as described in the section 3.2.3.

**Table S21.** Effect (HR) of smoking cessation to CHD and stroke event risk[58].

| | Men | Women |
|---|---|---|
| **CHD** | 0.66 | 0.55 |
| **Stroke** | 0.75 | 0.67 |

Abbreviations: CHD, coronary heart disease; HR, hazard ratio.

##### 3.3.2 Length of the risk reduction

The length of the risk reduction in T2D gained by weight loss was analyzed in a previous Finnish study[57]. Magnitude of the risk reduction revealed by the long-term follow-up was approximately the same as it was just after the intervention even after at least 15 years although the difference in weight between the intervention and control groups achieved through the intervention had decreased during the follow-up. However, the weight in the intervention group was 1% smaller compared with baseline still at year 10 of the follow-up and this may contribute to the long-lasting effect. In the present model, it was assumed that the risk reduction of T2D from weight loss remains similar for 15 years.

For risk reductions in other conditions the length of the reduction had to be assumed. With no better knowledge, the length was deemed to be analogous with the weight regain. There is not a collective result about the time nor the trajectory of weight regain after weight loss intervention, as the studies are heterogenous with methods, interventions and populations used and short of duration[59,60]. However, based on a published meta-analysis of nutritional weight loss interventions[60], the reduction was assumed to linearly diminish and reach the level of the normal development in 5 years.

#### 4 Costs

All the costs were adjusted to the level of 2022 according to the official price index of public expenditure and Index of wage and salary earnings[61,62]. Half-cycle correction and discount rate of 3% were used for costs in the model.

##### 4.1 Costs of health care service use

Cost of nurse health check, common laboratory tests connected to it, and physician's consultation were calculated with the costs related to them in public and private health care weighted with the proportion of the health checks within the provider[49]. The costs were estimated with rates of occupational health care within private providers and health promotion visits in public health care (**Table S22**). As time spent in the screening and preventive measures was reported as a measure of the resources needed, assumptions of 1 hour, 0.5 hour, and 10 minutes of working time from nurse, physician and laboratory, respectively, was spent per one individual using the service. For both nurse and physician visits, the estimated time included the full scope of the appointment: the reception, patient consultation and counselling, as well as documentation and other related administrative tasks[63].

**Table S22.** Costs and their references of the nurse health check, physician's consultation and laboratory tests.

| | Nurse health check (60 min) | Health check laboratory tests | Physician's consultation | Nurse's consultation (20 min) |
|---|---|---|---|---|
| **Occupational health nurse visits private occupational health services (incl. Kanta fee)** | 121.70€[1] | 159.0€[1] | | 44.47€[2] |
| **Public sector occupational health visit** | 64.00€[3] | 170.00€[3,4] | | 47.63€[5] |
| **20-minute physician consultation in private occupational health services (incl. Kanta fee)** | | | 74.00€[1] | |
| **Physician's consultation in public sector** | | | 93.00€[3] | |
| **Weighted average price (SD)** | 115.96€[6] (14.50) | 160.12€[6] (20.02) | 75.89€[6] (9.49) | 44.78€[6] |

[1] references [64–66]
[2] references [67–70]
[3] references [62,71]
[4] Calculated based on the average number of laboratory tests in private sector occupational health checkups and unit costs of a single laboratory test in public health care services.
[5] references [62,72]
[6] Calculated as a weighted average based on customer shares (private occupational health services 90.1% and public health services 9.9%)

The cost of MRS testing was obtained from the technology provider and set at €30 per individual. MRS testing was estimated to require 10 minutes of laboratory professional time per individual.

#### 4.2 Costs of preventive actions

The medication costs for primary prevention were the costs of antihypertensive and/or statin use described earlier. Additionally, for primary prevention, the cost of a control visit to physician's and nurse's office for the first year, and half of it for the following years (visits assumed for every other year) was added for modest approach, according to the Finnish standard health care costs[71] (**Table S23**). The control visit costs were added only once even if both, antihypertensive and cholesterol medication were used.

**Table S23.** Total costs of primary prevention.

| Primary prevention | Medication cost €/year (SD) | 1st year control visit costs € (SD) | Following years control visit costs €/year (SD) |
|---|---|---|---|
| **Antihypertensive medication** | 77 (9.6) | 132 (16.5) | 66 (8.2) |
| **Statins** | 28 (3.5) | 132 (16.5) | 66 (8.2) |

For smoking cessation among smokers, price on June 1st, 2022, of 12-week varenicline treatment was modelled as cost without the value tax. In Finland, varenicline (Champix) is sold in two packages, one for the starters with 0.5 mg and 1 mg doses (11 + 42 fol) and one for the continuers with 1 mg doses (112 fol) only. The treatment consists of one package each. It was assumed that all smokers that initiated the treatment also bought both packages. Thus, total cost of the smoking cessation in the model was 265€ (SD 33.8). In addition, one visit to physician's office was added to the expenses (93€).

The cost of Healthy weight coaching lifestyle intervention, 719€ (SD 92€) was defined as reported in previously published study and indexed to the year 2022[62,73].

#### 4.3 Costs of type 2 diabetes

The costs of T2D (**Table S24**) were modelled as in the previous diabetes model[74,75], except for medication costs. In summary, the costs include the additional costs for health care and productivity losses caused by diabetes and its complications retrieved from register studies[76–79] and data from a previous Finnish study described earlier[17]. As in previous studies, a variation of ±25% was applied to the costs where confidence interval was not reported. Medication costs were calculated from the Dispensations reimbursed under the NHI scheme register[32] and based on the costs of reimbursed purchases (value tax of 10% removed) and the number of recipients from the year 2022.

**Table S24**. The indexed costs of diabetes applied in the model.

| Cost parameter | Value, € (SD) | Reference |
|---|---|---|
| **Costs from productivity losses due to T2D** * | 9289 (1161) | [79] |
| **Cost of T2D complications**** | 13,201 (1650) | [80] |
| **Additional health care costs of T2D excluding primary health care**** | 3924 (490) | [77] |
| **Additional T2D health care costs for primary health care** | 621 (637) for men<br>599 (703) for women | [81] |
| **Additional medication costs of T2D**** | 567 (71) | [32] |

Abbreviations: T2D, type 2 diabetes.
*For persons under 65 years old.
**For variables without available confidence interval, a variation of ± 25% has been used as an estimate. In these cases, SE was calculated as: SE = (upper bound – lower bound)/ (1.96 x 2).

#### 4.4 Costs of CVD events

The costs of initial CHD event and stroke were modelled based on real-world study by *Sund et al, 2022*[80]. In the study the costs were estimated according to costs from the Hospital District of Northern Savo (KUH) and include the costs of the hospital treatment (inpatient days, surgical procedures, emergency department visits, outpatient physician and nurse visits, laboratory examinations, medications used in hospital and other accountable services related to treatment) of patients with T2D for the year following an initial event (**Table S25**). The difference of the costs of nondiabetics and diabetics was estimated to be only minor as, e.g., over half of CHD and stroke patients without diabetes might have abnormal blood glucose regulation[25,82]. Some bias can be caused by the differences of the definitions of the events as the diagnosis codes used in [80] are wider than the ones used in the actual event modelling, i.e., FINRISK 2.0. However, other studies have given very similar results on costs of CHD-events[83,84]. The cost of stroke is smaller than in older studies which can be due to the difference in diagnosis codes of the studies involved, e.g., treatment of TIA-attacks is included in the cost calculations, recent development in treatment of stroke, or both[85,86].

**Table S25.** Definitions of considered CVD events and related average cost estimates[80].

| | ICD-10 and procedure codes | Cost, € | SD |
|---|---|---|---|
| **CHD event** | I20–I25, I46, I48, I50, R96, R98, FNA-FNE, FN1AT, FN1BT, FN1ST, FN1YT, FN2, FN3CT, TFN40, TFN50 | 14,224 | 20,728 |
| **Stroke** | I63.0–I63.5, I63.7–I66.9, I67.2, I69.3, G45 | 13,815 | 21,013 |

Abbreviations: CHD, coronary heart disease.

The costs of rehabilitation and productivity losses were added to the cost of the treatment for non-fatal CHD-events and strokes. The total costs of rehabilitation used for ischaemic heart disease and cerebrovascular events, according to ICD-10 codes, were retrieved from SII[87,88] and divided evenly for every patient in rehabilitation with the same diagnosis (**Table S26**). The probability to go to rehabilitation was estimated by the proportion of patients in rehabilitation of all the incident CHD or stroke events of the same year from PERFECT[48] (**Table S26**).

**Table S26.** Cost and probability of rehabilitation.

| Event diagnosis | Year | Total costs, 1,000€ | Number of patients in rehabilitation | Cost, €/patient (SD) | Number of new patients | Proportion of patients in rehabilitation (%) |
|---|---|---|---|---|---|---|
| **I20-I25** | 2019 | 1,622 | 912 | 1,779* (222) | 11,311 | 8.0 |
| **I60-I69** | 2018 | 19,696 | 2,617 | 7,527* (941) | 9,668 | 27.1 |

*For variables without available confidence interval, a variation of ± 25% has been used as an estimate. In these cases, SE was calculated as: SE = (upper bound – lower bound)/ (1.96 x 2).

Productivity losses for the year of the event were calculated according to average length of sick leave after CHD or stroke event (in weekdays)[89] and average cost of one sickness absence workday (344€) in Finland in 2022[90] respectively (**Table S27**). In Finland, one week typically contains 5 workdays. Average length of sick leave reported in Statistical database Kelasto contains only weekdays (6 days per week) and does not include the first 10 days of the sick leave that are not reimbursed by the Social Insurance Institute of Finland. Therefore, weekdays were adjusted to cover also the 10 first sickness absence weekdays and thereafter the number was converted to workdays by multiplying the sick leave by the 5 out of 6. The productivity losses were added to the total costs of people under 65 years and surviving the event.

**Table S27.** The productivity losses for the year of the event.

| Event diagnosis | Average length of sick leave, workdays | Average cost of sickness absence, €/workday | Productivity loss after event, €/patient (SD) |
|---|---|---|---|
| **I20–I22** | 52.2 | 344 | 17,957 (2290) |
| **I61, I63-I64** | 126.8 | 344 | 43,619 (5564) |

Productivity losses for the following years were added also for patients under 65 years old that were unable to return to work after CVD-event. The proportion of patients going on disability pension was calculated separately for CHD and stroke events according to the number of initiated disability pensions by diagnosis from the SII, and the number of patients under 65 years old surviving the event, both from year 2014[5,6] (**Table S28**). Productivity losses were calculated for every year, excluding the year of the event, until individual turned 65 or until death with 12.5*average monthly wage.

**Table S28.** Proportion of patients on disability pension and the annual productivity loss per patient.

| | Proportion developing disability to work (%) | Yearly cost of the disability (indexed), €/patient (SD) |
|---|---|---|
| **CHD** | 4.7 | 47,151 (5894) |
| **Stroke** | 24.2 | 47,151 (5894) |

Death from ACS or stroke was determined not to cause any costs. This is not accurate for all deaths even though major part of ACS deaths takes place before admission to a hospital[24]. This approach was selected for simplicity.

Costs of CHD and stroke after the first event were calculated as average yearly costs caused by medication and estimated need for health care visits based on the Finnish Current Care Guidelines[24,25]. Condition related compilation of treatment and its costs is described in **Table S29**. As the incidence of subsequent events was not modelled, the costs of the treatment were not included in the model. Thus, the costs are an underestimation of the actual costs.

**Table S29.** Annual costs of the conditions per patient after the initial event.

| Condition | Medication | Healthcare visits /year | Total costs, €/year (SD) |
|---|---|---|---|
| **CHD** | antihypertensive<br>lipid-lowering<br>antithrombotic | 1 * nurse 39€<br>1 * physician 93€ | 107+30+39+93=269 (34) |
| **Stroke** | For 86.5% of the patients:<br>antihypertensive<br>lipid-lowering<br>antithrombotic<br>For 13.5% of the patients:<br>antihypertensive | 1 * nurse 39€<br><br><br><br>1 * nurse 39€ | 0.865*(296+28+77+39)<br>+ 0.135*(77+39) = 396 (51) |

Abbreviations: CHD, coronary heart disease.

The cost of medication after CHD event was calculated from Dispensations reimbursed under the NHI scheme register[32] by calculating the average per patient cost of medication purchases reimbursed with the special imbursement number for CHD (206) in the year 2022. The costs were calculated to be 107€/year (value tax removed). As small dose (100 mg) acetylsalicylic acid (ASA) is part of regular drug therapy after CHD event, but it is not reimbursable, it was added to the costs separately. As small dose ASA does not have a preference price, the price was calculated using the most affordable price in 1–14 November 2023 and assuming 100% adherence (30€/year, value tax removed). The costs of one visit to physician's (indexed á 93€) and nurse's (á 39€) office in primary health care per year was added to medication expenses[62,71]. The visit costs also include the possible laboratory expenses.

As there are notable differences in the treatment and secondary prevention of intracranial haemorrhage (ICH) and ischaemic stroke (IS), the weighted average based on the proportion of ICH and IS survivors, according to event case numbers and their case fatalities from the year 2014[5] was used to evaluate the costs after stroke event. The costs for antihypertensive medication for ICH and antihypertensive, lipid-lowering and antithrombotic medication for IS were weighted with 13.5% and 86.5%, respectively. For simplicity, a modest estimation of one visit to the nurse's office per year was added as follow-up cost to both ICH and IS.

For antithrombotic medication costs after IS the yearly cost of the combination of ASA and dipyridamole (25mg/200mg x2) was added, as it is the first option for secondary prevention in the Current Care Guidelines in Finland[25]. The cost was calculated to be 296€/year (based on the price for 1-14 November 2023, value tax removed).

The yearly costs of antihypertensive medication and statins were retrieved from Dispensations reimbursed under the NHI scheme register[32]. The mean price per statin or antihypertensive medication user was calculated using the statistics from 2022. The prices were concluded to be 28€ and 77€ (value tax removed) per year per user for statins and antihypertensive medication, respectively. The prices contain some bias as the numbers include both, primary and secondary prevention users (e.g., different strengths) and users for other indications (e.g., heart failure or atrial fibrillation). Additionally, there are users that die or otherwise stop the medication use in the middle of the year, causing them to buy less of the drug, and their presence causes an underestimation of the mean price of the full year adherent users. The price is, however, a tolerable estimate of costs of a user with an average adherence excluding only users who do not purchase the medicine at all (i.e., adherence is 0%).

#### 5 Utilities

The utility values used in this model were all based on EQ-5D-3L measuring instrument. Baseline utilities and disutility due to diabetes and its complications were modelled as in several previous diabetes-models[74,75,91] (**Table S30**). Baseline utilities depended on age and sex. Disutility because of diabetes complications was a prevalence-weighted mean of disutility of each complication. The utility values and disutility because of diabetes were based on studies from Finland and disutility of complications was calculated on basis of prevalence of complications from Finland and disutility values from other parts of Europe as described in *Martikainen et al., 2022*[75].Disutility of the year of the incident CVD event was modelled according to a Swedish sub-study of Anglo-Scandinavian cardiac outcomes trial (ASCOT)[92] with 60 patients with initial CVD-event and their nearly 400 controls (**Table S30**). Disutility values after the first year were modelled with disutility values from a Finnish survey study with 8028 participants where the disutility connected to prevalent conditions was studied[93]. Half-cycle correction and discount rate of 3% were used for utilities in the model. The potential positive effects of intervention, i.e., weight loss, on quality of life were not modelled.

**Table S30.** Utility parameters applied for baseline utility and disutility because of T2D in the model, and their distributions.

| Utility | Value and variation | Distribution | Reference |
|---|---|---|---|
| **Baseline utilities:** | | Beta | 91 |
| **Women (SE)** | | | |
| **30–44** | 0.906 (0.003) | | |
| **45–54** | 0.865 (0.005) | | |
| **55–64** | 0.810 (0.006) | | |
| **65–** | 0.770 (0.008) | | |
| **Men (SE)** | | | |
| **30–44** | 0.917 (0.003) | | |
| **45–54** | 0.876 (0.005) | | |
| **55–64** | 0.821 (0.006) | | |
| **65–** | 0.781 (0.008) | | |
| **Disutility of T2D (SE)** | 0.041 (0.012) | Beta | 93 |
| **Disutility of T2D complications** | 0.119 (0.078-0.160) | Beta | 75 |
| **Disutility of CHD** | | Beta | |
| **First year (SD)** | 0.051 (0.126) | | 92 |
| **Following years (SE)** | 0.011 (0.009) | | 93 |
| **Disutility of stroke** | | Beta | 77 |
| **First year (SD)** | 0.145 (0.145) | | 92 |
| **Following years (SE)** | 0.090 (0.020) | | 93 |

Abbreviations: CHD, coronary heart disease; T2D, type 2 diabetes.

#### 6 Validation

The model was reviewed and validated along construction by exploring the distributions of the simulated baseline characteristics, incidence of preventive actions (such as initiation of statins and antihypertensive medication, smoking cessation intervention, healthy weight coaching, and visits to nurses and physicians), incidence of CVD and T2D events as well as costs by scenarios and risk categories from the simulation results obtained. Distributions of baseline characteristics and incidence of CVD and T2D events were compared with those of the observed data. The model was iteratively updated when errors were detected.

#### 7 Sensitivity analyses

In addition to primary analyses, sensitivity analyses against time consumption were investigated. Two sensitivity analyses were carried out. First, attendance rate equivalent to that of sHC (i.e., 76.0% over five years) was utilized also for mHC. The results from the base-case analysis resembled those of the primary analyses with mHC being a dominant option (i.e., more effective and less costly) with a modest reduction in gained QALYs due to delayed initiation of preventive actions and €2.4 million higher savings obtained in the target population (**Tables S30–S31**).

Second, a nurse visit of 20 minutes was added to the moderate- and high-risk individuals in the mHC to reflect the current practise used in Finnish healthcare. In the base-case scenario, nurse work time savings decreased from 120.7 full-time equivalent (FTE) years to 67.8 FTE years together with an overall reduction of €3.6 million in the savings in the target population when sHC was compared against mHC (**Tables S32–S33**). Still, the mHC was a dominant option (i.e., more effective and less costly).

**Table S31.** Time required for risk identification and associated FTE gains from an additional analysis assuming equivalent attendance rate in both the standard and MRS-based health checks (i.e., 76.0% over five years). Time required to identify individuals at risk in a 50–54-year-old population (n=256,372), comparing standard and MRS-based health checks. Values represent total hours for nurses, physicians, and laboratory work over five years, and corresponding full-time equivalent (FTE) working years gained. One FTE equals 1,600 hours. In the standard health check, risk identification includes laboratory testing (10 min) and a nurse visit (1 h), with a subsequent physician visit (30 min) for high-risk individuals. In the MRS-based health check, risk identification is based on laboratory testing (10 min), followed by a physician visit (30 min) for high-risk individuals.

| Arm | Nurse (h) | Physician (h) | Laboratory (h) | Gained nurse FTE years | Gained physician FTE years |
|---|---|---|---|---|---|
| **Standard health check** | 193,377 | 14,758 | 32,230 | | |
| **MRS-based health check** | 0 | 12,032 | 32,474 | | |
| **Difference between MRS-based and standard health check** | -193,377 | -2,727 | 244 | 120.9 | 1.7 |

Abbreviations: FTE, full-time equivalent; MRS, metabolomic risk score.

**Table S32.** Incremental costs and QALYs of MRS-based health check versus standard health check from an additional analysis assuming equivalent attendance rate in both the standard and MRS-based health checks (i.e., 76.0% over five years). Savings, gained QALYs and ICER/QALY in the target population of 50–54-year-old individuals in labour force (n=256,372).

| | Life years per individual | QALYs, per individual | Total costs per individual, € | Difference in QALYs per individual | Gained QALYs in the target population | Difference in costs per individual, € | Savings in the target population, € | ICER/QALY, € |
|---|---|---|---|---|---|---|---|---|
| **Base-case analysis** | | | | | | | | |
| Standard health check | 31.8 | 15.60 | 15,399 | | | | | |
| MRS-based health check | 31.8 | 15.61 | 15,286 | 0.01 | 1,123 | -113 | -28,963,371 | Dominant |
| **Scenario analysis 2** | | | | | | | | |
| Standard health check | 31.8 | 15.60 | 15,399 | | | | | |
| MRS-based health check | 31.9 | 15.64 | 14,437 | 0.04 | 11,028 | -962 | -246,560,005 | Dominant |
| **Scenario analysis 3** | | | | | | | | |
| Standard health check | 31.9 | 15.65 | 14,544 | | | | | |
| MRS-based health check | 31.9 | 15.64 | 14,437 | -0.01 | -1,366 | -107 | -27,385,059 | NA |

Abbreviations: ICER, incremental cost-effectiveness ratio; MRS, metabolomic risk score; NA, not applicable; QALY, quality-adjusted life year.

**Table S33.** Time required for risk identification and associated FTE gains from an additional analysis adding a nurse visit of 20 minutes to the moderate- and high-risk individuals in the MRS-based health check. Time required to identify individuals at risk in a 50–54-year-old population (n=256,372), comparing standard and MRS-based health checks. Values represent total hours for nurses, physicians, and laboratory work over five years, and corresponding full-time equivalent (FTE) working years gained. One FTE equals 1,600 hours. In the standard health check, risk identification includes laboratory testing (10 min) and a nurse visit (1 h), with a subsequent physician visit (30 min) for high-risk individuals. In the MRS-based health check, risk identification is based on laboratory testing (10 min), followed by a nurse visit (20 min) for moderate- and high-risk individuals and a physician visit (30 min) for high-risk individuals.

| Arm | Nurse (h) | Physician (h) | Laboratory (h) | Gained nurse FTE years | Gained physician FTE years |
|---|---|---|---|---|---|
| **Standard health check** | 195,815 | 15,179 | 32,636 | | |
| **MRS-based health check** | 80,471 | 12,066 | 32,474 | | |
| **Difference between MRS-based and standard health check** | -115,344 | -3,113 | -162 | 67.8 | 1.9 |

Abbreviations: FTE, full-time equivalent; MRS, metabolomic risk score.

**Table S34.** Incremental costs and QALYs of MRS-based health check versus standard health check from sensitivity analyses adding a nurse visit of 20 minutes to the moderate- and high-risk individuals in the MRS-based health check. Savings, gained QALYs and ICER/QALY in the target population of 50–54-year-old individuals in labour force (n=256,372).

| | Life years per individual | QALYs, per individual | Total costs per individual, € | Difference in QALYs per individual | Gained QALYs in the target population | Difference in costs per individual, € | Savings in the target population, € | ICER/QALY, € |
|---|---|---|---|---|---|---|---|---|
| **Base-case analysis** | | | | | | | | |
| Standard health check | 31.8 | 15.60 | 15,453 | | | | | |
| MRS-based health check | 31.8 | 15.60 | 15,350 | 0.01 | 1,316 | -90 | -22,963,593 | Dominant |
| **Scenario analysis 2** | | | | | | | | |
| Standard health check | 31.8 | 15.60 | 15,453 | | | | | |
| MRS-based health check | 31.9 | 15.64 | 14,448 | 0.04 | 10,813 | -992 | -254,199,900 | Dominant |
| **Scenario analysis 3** | | | | | | | | |
| Standard health check | 31.9 | 15.65 | 14,590 | | | | | |
| MRS-based health check | 31.9 | 15.64 | 14,448 | -0.01 | -1,672 | -128 | -32,750,997 | NA |

Abbreviations: ICER, incremental cost-effectiveness ratio; MRS, metabolomic risk score; NA, not applicable; QALY, quality-adjusted life year.
